# Genetic Contributions to Early and Late Onset Ischemic Stroke

**DOI:** 10.1101/2021.11.06.21265795

**Authors:** Thomas Jaworek, Huichun Xu, Brady J. Gaynor, John W. Cole, Kristiina Rannikmäe, Tara M. Stanne, Liisa Tomppo, Vida Abedi, Philippe Amouyel, Nicole. D Armstrong, John Attia, Steven Bell, Oscar R. Benavente, Giorgio B. Boncoraglio, Adam Butterworth, the Cervical Artery Dissections and Ischemic Stroke Patients (CADISP) Consortium, Jara Cárcel-Márquez, Zhengming Chen, Michael Chong, Carlos Cruchaga, Mary Cushman, John Danesh, Stephanie Debette, David J. Duggan, Jon Peter Durda, Gunnar Engstrom, Christian Enzinger, Jessica D. Faul, Natalie S. Fecteau, Israel Fernández-Cadenas, Christian Geiger, Anne-Katrin Giese, Raji P. Grewal, Ulrike Grittner, Aki S. Havulinna, Laura Heitsch, Marc C. Hochberg, Elizabeth Holliday, Jie Hu, Andreea Ilinca, the International Stroke Genetics Consortium (ISGC), the INVENT Consortium, Marguerite R. Irvin, Rebecca D. Jackson, Mina A. Jacob, Raquel Rabionet Janssen, Jordi Jimenez-Conde, Julie A. Johnson, Yoichiro Kamatani, Sharon L.R. Kardia, Masaru Koido, Michiaki Kubo, Leslie Lange, Jin-Moo Lee, Robin Lemmens, Christopher Levi, Jiang Li, Liming Li, Kuang Lin, Haley Lopez, Sothear Luke, Jane Maguire, Patrick F. McArdle, Caitrin W. McDonough, James F. Meschia, Tiina Metso, Martina Müller-Nurasyid, Timothy O’Connor, Martin O’Donnell, Leema Reddy Peddareddygari, Joanna Pera, James A. Perry, Annette Peters, Jukka Putaala, Debashree Ray, Kathryn Rexrode, Marta Ribases, Jonathan Rosand, Peter M. Rothwell, Tatjana Rundek, Kathleen A. Ryan, Ralph L. Sacco, Veikko Salomaa, Cristina Sanchez-Mora, Reinhold Schmidt, Pankaj Sharma, Agnieszka Slowik, Jennifer A. Smith, Nicholas L. Smith, Sylvia Wassertheil-Smoller, Martin Soderholm, O. Colin Stine, Daniel Strbian, Cathie LM Sudlow, Turgut Tatlisumak, Chikashi Terao, Vincent Thijs, Nuria P. Torres-Aguila, David-Alexandre Trégouët, Anil Man Tuladhar, Jan H. Veldink, Robin G. Walters, David R. Weir, Daniel Woo, Bradford B. Worrall, Charles C Hong, Owen Ross, Ramin Zand, F-E de Leeuw, Arne G. Lindgren, Guillaume Pare, Christopher D. Anderson, Hugh S. Markus, Christina Jern, Rainer Malik, Martin Dichgans, Braxton D. Mitchell, Steven J. Kittner, International Stroke Genetics Consortium, Early Onset Stroke Genetics Consortium

## Abstract

**Objective:** To determine the contribution of common genetic variants to risk of early onset ischemic stroke (IS).

**Methods:** We performed a meta-analysis of genome-wide association studies of early onset IS, ages 18-59, using individual level data or summary statistics in 16,927 cases and 576,353 non-stroke controls from 48 different studies across North America, Europe, and Asia. We further compared effect sizes at our most genome-wide significant loci between early and late onset IS and compared polygenic risk scores for venous thromboembolism between early versus later onset IS.

**Results:** We observed an association between early onset IS and *ABO*, a known stroke locus. The effect size of the peak ABO SNP, rs8176685, was significantly larger in early compared to late onset IS (OR 1.17 (95% C.I.: 1.11-1.22) vs 1.05 (0.99-1.12); p for interaction = 0.008). Analysis of genetically determined ABO blood groups revealed that early onset IS cases were more likely to have blood group A and less likely to have blood group O compared to both non-stroke controls and to late onset IS cases. Using polygenic risk scores, we observed that greater genetic risk for venous thromboembolism, another prothrombotic condition, was more strongly associated with early, compared to late, onset IS (p=0.008).

**Conclusion:** The *ABO* locus, genetically predicted blood group A, and higher genetic propensity for venous thrombosis are more strongly associated with early onset IS, compared with late onset IS, supporting a stronger role of prothrombotic factors in early onset IS.

## Introduction

Substantial advances have been made in recent years towards identifying common genetic variation associated with risk of ischemic stroke (IS) ^1, 2^. This progress has been largely based on meta-analysis of genome-wide association study (GWAS) results derived from predominantly late onset cases. Given that a higher heritability of early onset IS observed in multiple studies^3-6^, there is a strong need for genetics studies focusing on early onset stroke. A pressing question is whether the genetic contribution to early onset stroke includes mechanisms that may be novel or specific to early onset ischemic stroke but may still have translational importance across the whole age spectrum as has been found from studies of early onset cases in other complex diseases ^7-9^.

Because atherosclerosis is a less common cause of stroke in young adults, we hypothesized that non-atherosclerotic, prothrombotic mechanisms may be more important and discernable in studies of early onset IS ^10, 11^. This concept is supported by associations reported between early onset IS and multiple prothrombotic candidate genes ^10-14^. In this report, we present findings from the Genetics of Early Onset Ischemic Stroke Consortium, contrast the effect sizes of known stroke loci in early versus later onset stroke, and evaluate differing contributions of prothrombotic loci to early and late onset IS.

## Methods

The Early Onset Stroke Consortium (EOSC) is a collaboration of investigators representing 48 different studies across North America, Europe, Japan, Pakistan, and Australia who have pooled their data for a GWAS meta-analysis of early onset IS in cases aged 18-59 years. Collectively, these studies contributed 17,077 cases (16,927 cases included for analysis) and 576,353 non-stroke controls (eMethods and eTable 1). All patients had brain imaging to exclude diagnoses other than IS. Additional screening was performed in some, but not all, studies to exclude cases believed to be due to a known monogenic cause (e.g., sickle cell disease) or to a known non-genetic cause (e.g., drug use, complications of procedures). Ischemic stroke subtyping was performed using the TOAST criteria ^15^ by most, but not all, sites.

The EOSC includes cases from two different sources: early onset stroke cases who previously participated in the Stroke Genetics Network (SiGN) ^16^ (n = 7,619), and early onset cases from additional non-SiGN study sites (n = 9,598). eTable 1 lists the 48 sites contributing early onset stroke cases and sources of controls. With one exception (Group 7 including cases from Barcelona and BASICMAR), controls from each study were of the same age or older than cases. Analysis groups were assigned as previously described to combine cases and controls of similar genetic ancestry groups and genotyped on arrays of similar density ^16^. Clinical characteristics of the stroke cases are shown in eTable 2. Genotypes for all studies except Helsinki were imputed using the TOPMed reference panel on the University of Michigan Imputation Server ^17^. The Helsinki Study imputed genotypes using a Finnish population-specific reference panel and the BEAGLE software. All genotype data were based on genome build hg38. Genotyping platforms and cohort-specific quality control and analysis parameters are provided in eTable 3.

GWAS of stroke cases and controls were conducted within sites or within groupings of sites, and then meta-analyzed. Prior to analysis, we removed stroke cases from analysis if there were fewer than 40 in the analysis strata, if they could not be assigned to a genetic ancestry group, or if there was not an adequate number of controls. Filtering on these criteria left 16,927 (of 17,077) early onset cases for analysis. The overall analytic design is depicted in Figure 1. Our primary analysis was a transethnic meta-analysis. In parallel, we performed a European only meta-analysis. Logistic regression was performed to test for association between stroke occurrence and single variants. Covariates included sex and up to 10 principal components to adjust for population stratification. Power calculations indicated that our study provided 80% power to detect odds ratios ranging from 1.09 to 1.20 for common genetic variants with minor allele frequencies (MAF) > 5% at the genome-wide threshold for significance, i.e., 5 × 10^−8^. For comparison, the previously detected odds ratios for IS from MEGASTROKE GWAS (67,162 cases) ranged from 1.05 – 1.09 ^1^. We also performed TOAST-defined stroke subtype analyses for those sites providing subtype classification.

**Figure 1.**
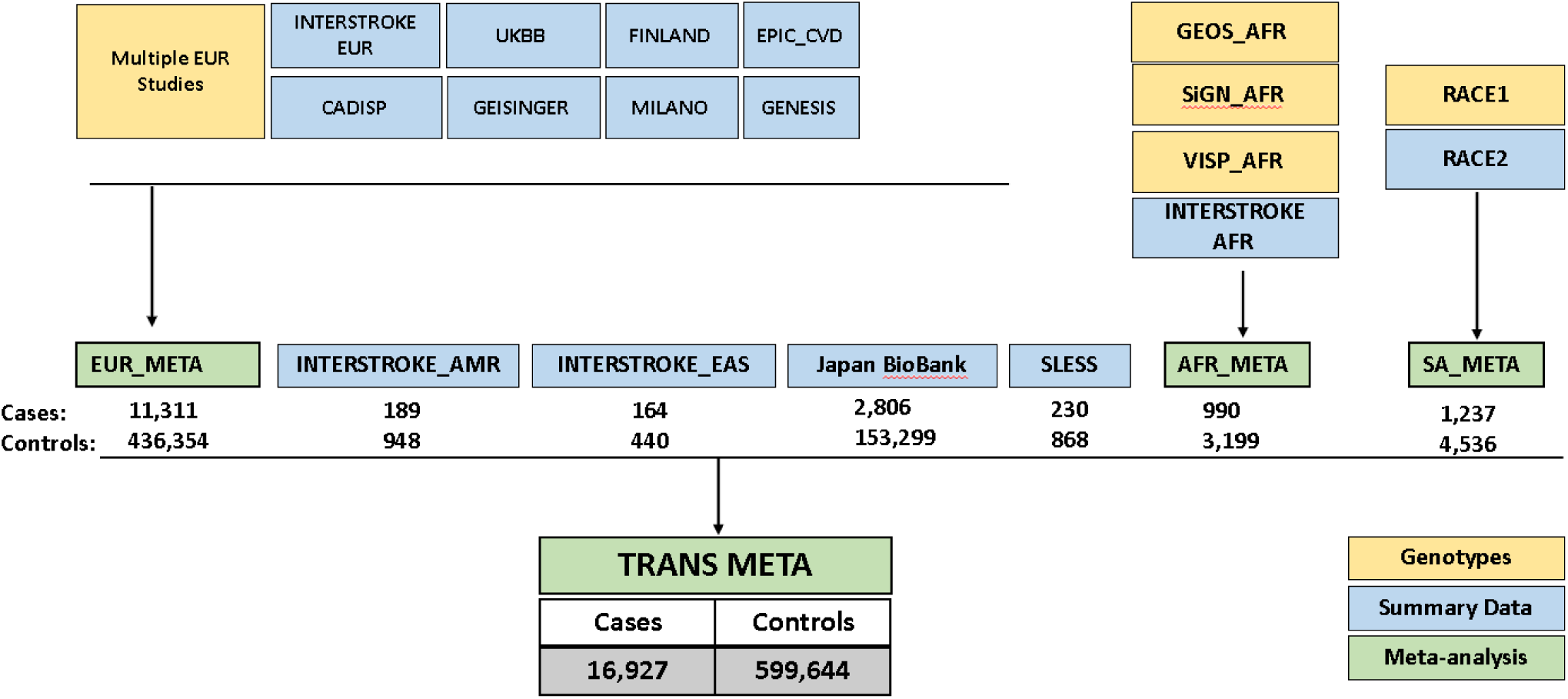
Overview of Study Design *Case/Control number equals the number included in the final analysis. **See eTable 1 for list of the studies with genotype data used in the European meta-analysis

We compared effect sizes between early and late onset stroke cases of European ancestry at 40 loci previously associated with IS in MEGASTROKE, ^1^ MEGASTROKE and UK Biobank (UKB) combined, ^2^ or in a previously published meta-analysis of small vessel stroke ^18^. A late onset stroke case cohort (age onset ≥ 60 years) from the SiGN Consortium consisting of 9,272 late onset cases and 25,124 controls of European ancestry was created for this comparison. Effect sizes between early and late onset stroke were compared using a Wald test.

To follow-up our peak association at the *ABO* locus, we assessed the association of early onset stroke with *ABO* blood groups and then compared these associations with late onset stroke. The *ABO* locus encodes two glycosyltransferases, A and B, that define the serologic blood groups, A, B, AB, and O. To explore associations of ABO blood group with stroke, we first tested whether histo-blood group ABO was associated with IS by comparing the distribution of these blood groups among early and late onset cases and controls. We assigned the diploid ABO blood group using genotypes at two SNPs (rs8176719 and rs8176746), as described by Groot et al. ^19^ (see eMethods).

At the haploid level, the ABO blood groups and subgroups are defined by 5 common haplotypes, each of which is tagged by a single SNP ^20^ (see eMethods). The 5 *ABO* haplotypes are A1 (the ancestral haplotype, tagged by rs2519093-T), O1 (tagged by a frameshift deletion, rs8176719-delG), A2 (tagged by rs1053878-A), B (tagged by rs8176743-T), and O2 (tagged by rs41302905-T)^20^. Goumidi and colleagues have recently shown that relative to haplotype O1, the genetically defined haplotypes A1 and B are strongly associated with venous thrombosis risk, and haplotype A2 is associated with a modest increase in risk ^20^.

We hypothesized that the strong association of early onset stroke with the *ABO* locus was related to the prothrombotic properties of the ABO blood group. We evaluated this hypothesis first by testing whether the two lead *ABO* variants were associated with venous thromboembolism (VTE), another prothrombotic condition, in the UKB and if the association was more prominent in early compared to later onset VTE. Using summary level association results (VTE results from the INVENT Consortium ^21^), we then estimated pairwise genetic correlations among early onset stroke, late onset stroke, and VTE using LD Score Regression analysis (LDSC) ^22^. We then tested if genetic predisposition to VTE, as measured by a polygenic risk score (PRS), was more prominently associated with early compared to later onset stroke. For this purpose, we generated a VTE PRS for individuals from the EOSC and late onset stroke subset from SiGN based on a large prior GWAS of VTE ^23^ using PRSice software ^24^. The VTE PRS included 255 SNPs using a GWAS p-value threshold of 1 × 10^™5^ (see eMethods). We tested the association between the VTE PRS score with stroke in the European ancestry sample using logistic regression with 10 principal components for ancestry and sex included as covariates. Effect sizes between early and late onset stroke were compared using a Wald test (See eMethods).

We identified several disorders and plasma biomarkers (late onset stroke, VTE, and plasma levels of von Willebrand factor (VWF) and Factor VIII) for which associations at the ABO locus have previously been reported and then used the coloc software ^25^ to assess evidence that the same causal SNPs associated with EOS also drove associations with the second trait. Briefly, coloc utilizes a Bayesian approach and summary level association level results for two traits to calculate the posterior probabilities of five competing hypotheses (H0-H4) that assess whether the associations are due to the same (corresponding to H4) or a different (corresponding to H3) causal variant (see eMethods).

### Standard Protocol Approvals, Registrations, and Patient Consents

All participating sites obtained IRB or Ethics Board approval, and informed consent was obtained from all participants or their legally authorized representative.

### Data Availability

Summary results will be made available upon application to the contact authors and consortium approval of the request. Individual level data from a subset of sites will be made available on dbGaP.

## Results

### Genome-wide analysis results

The trans-ethnic meta-analysis included a total of 16,927 early onset IS cases, representing 1,073 of African ancestry,10,549 of European ancestry, 189 of Hispanic ancestry, 230 of Afro-Caribbean ancestry and 4,204 of pan-Asian ancestry. There was little evidence for inflation of p-values across sites, as indicated by genomic control ranging from 0.86 to 1.14 (eTable 3). We identified two loci associated with early onset IS at genome-wide significance. The most strongly associated variant at the first locus was rs8176685 with an odds ratio (OR) of 1.17 (95% confidence interval [95% CI]: 1.11-1.22; p = 8.25 × 10^−11^), marking a 12 base pair insertion/deletion polymorphism at the *ABO* locus, a locus previously associated with IS ^1, 26^ (Figure 1A and eTable 4), and in near perfect linkage disequilibrium with rs635634, the lead *ABO* SNP associated with all IS in MEGASTROKE ^1^.

The second locus significantly associated with early onset IS was *MLXIP* (MLX Interacting Protein) (rs377424471, OR=0.90, 95% CI: 0.86-0.94; p=3.96 × 10^−8^) (eTable 4). To our knowledge, this locus has not previously been associated with stroke. This SNP showed no evidence for association with late onset stroke in SiGN (OR = 1.00, 95% CI: 0.97-1.04, p = 0.99), further highlighting the need for this result to be replicated.

The European ancestry only meta-analysis, based on 10,549 early onset stroke cases, also indicated a strong association with the *ABO* locus, albeit with a different lead SNP, rs529565 (previously rs912805253). *ABO* rs529565 T allele had an OR for association of 0.88 (95% CI: 0.85-0.91; p = 3.17 × 10^−13^) (Figure 1B and eTable 5). Stratified analyses revealed no difference in effect sizes of either *ABO* rs8176685 or rs529565 between men and women.

In addition to the two genome-wide significant loci, we observed 23 “suggestive” loci with sub-threshold levels of significance (i.e., p < 1 × 10^−6^) in the trans-ethnic analysis (eTable 4) and 15 loci in the European only analysis (eTable 5) (9 of which were EUR only). Among these loci were *SH2B3* (SH2B Adaptor Protein 3, rs3184504), which has previously been associated with IS ^1^, and SNPs at several compelling candidate loci, including *F11* (Plasma Thromboplastin Antecedent) encoding factor XI in the coagulation cascade, and *MC4R* (Melanocortin Receptor 4), an obesity-associated gene. Twelve of the 32 unique loci associated with early onset IS in either the Transethnic or European only analysis at sub-genome thresholds (i.e., p<1 × 10^−6^) were nominally associated with late onset IS at p < 0.05 (eTables 4 and 5), and the odds ratios for all 12 were approximately similar between early and late onset stroke.

Analysis of IS subtypes revealed 11 SNPs associated with stroke subtypes at genome-wide thresholds of significance (eTable 6), although the sample sizes were relatively small for each subtype (ranging from 886-5149 for transethnic analysis and 376-1502 for European only analysis) and the frequencies of the associated SNPs were low (8 with EUR MAF < 0.02 and the 3 remaining MAF < 0.09).

There was no evidence for replication of the *HABP2* rs11196288 variant, which was previously associated with all early onset IS in our earlier phase 1 transethnic meta-analysis from the EOSC ^12^, although its minor allele frequency is only ∼3% in European ancestry populations, as indicated in gnomAD). In the expanded meta-analysis presented in this report (16,927 currently vs 4,505 cases previously), there was no evidence for association of this SNP with early onset stroke in any of the new sites, including those of non-European ancestry (eFigure 1).

### Associations of index ABO variants with all IS and IS subtypes in early and late onset IS

As described below, the rs8176685 deletion and the rs529565 T allele at the *ABO* locus tag blood groups A1 and O1, respectively. The associations we observed for these SNPs with early onset stroke are substantially higher than the peak associations previously reported at the *ABO* locus in predominantly older stroke populations (e.g., OR = 1.08; 95% CI: 1.05-1.11 in MEGASTROKE^1^). Both SNPs also had significantly larger effect sizes for all IS in early compared to late onset IS (*ABO* rs8176685: OR = 1.17 [95% CI: 1.11-1.22] vs 1.05 [95% CI: 0.99-1.12], and *ABO* rs529565: OR = 0.88 [95% CI: 0.85-0.92] vs 0.96 [95% CI: 0.91-1.00], p-values for interaction = 0.008 and 0.0006, respectively).

In subtype-specific analyses, the effect sizes of the A1-defining SNP rs8176685 did not differ significantly between early and late onset IS except for undetermined strokes, for which the A1-defining allele had a stronger effect in early compared to late onset stroke (p-value for homogeneity = 0.011). In contrast, the O1-defining allele at rs529565 was more protective in early versus late onset IS for large artery stroke, cardioembolic stroke, and undetermined stroke (p-values for homogeneity = 0.040, 0.040 and 0.002 respectively) (eFigures 2a and 2b).

We assessed the associations of *ABO* SNPs rs2519093 (in high LD with rs8176685, which was not present in UKB, and encoding blood group A1) and rs529565 (encoding blood group O1) with early and late onset IS in the UKB as a quasi-replication, ‘quasi’ because early onset UKB cases were included as part of the primary EOSC analyses, but late onset cases were not. The analysis was limited to IS cases and based on ICD codes, as described in the eMethods. Similar to our analysis in EOSC and SiGN, we observed stronger associations of both SNPs in early than in late onset stroke. The odds ratios of *ABO* rs2519093 (A1-defining) were 1.10 (95% CI: 1.01-1.19; p = 0.03) for early and 1.05 (95% CI: 1.00-1.02; p = 0.07) for late onset stroke, and the odds ratios of *ABO* rs529565 (O1-defining) were 0.93 (95% CI: 0.86-0.99; p = 0.02) for early and 0.95 (95% CI: 0.90-0.99; p = 0.02) for late onset stroke.

We performed a joint analysis of *ABO* SNPs rs8176685 and rs529565 to assess their independent associations with early onset IS. The frequencies of the rs8176685 deletion and the rs529565 T allele at the *ABO* locus in European ancestry populations are 0.20 and 0.64, respectively, leading to a modest correlation between them (r^2^=0.44), although the rs8176685 deletion appears only on the background of the more common rs529565 T allele, resulting in a D’ of 1 (eFigure 3). Because of the high D’ between these 2 SNPs, we used a stratified approach to assess the contribution of rs529565 to stroke risk in the absence of the rs8176685 deletion; that is, we estimated the association of rs529565 with stroke in individuals without the rs8176685 T allele. In this analysis, which was restricted to those strata for whom we had individual level data, rs529565 remained associated with all IS and at approximately the same effect size (OR = 0.90, 95% CI: 0.85-0.96; p=0.003), implying an association of this SNP that was independent of the rs8176685 indel SNP.

### Associations of ABO blood group-defining SNPs with early and late onset stroke

We tested associations of these blood group-defining haplotypes with early and late onset stroke in the European ancestry subgroup. As indicated previously, the top *ABO* SNPs identified in our GWAS, rs8176685 and rs529565, are in high linkage disequilibrium with the tagging SNPs for haplotypes A1 and O1, which were not included in our dataset. We also used rs1137827 as a high LD tag for rs8176743 (blood group B) (Table 1). Consistent with the VTE findings from Goumidi et al, our analyses indicate haplotype A1 (rs8176685) to be strongly associated with early onset stroke (OR = 1.18, 95% CI: 1.11-1.25; p = 1.95 × 10^−8^ and blood group O1 (rs529565) to be protective (OR = 0.88, 95% CI: 0.85-0.91; p = 3.17 × 10^−13^) (Table 1). Unlike for VTE, blood group B (rs1137827) showed little evidence for association with stroke (OR = 1.04, 95% CI: 0.94-1.14; p = 0.463. These trends persisted for late onset stroke, although the strength of associations was markedly reduced; that is, blood group A1 was only modestly associated with late onset stroke (OR = 1.05, 95% CI: 1.00-1.10; p = 0.044) and blood group O1 was only modestly associated with protection against late onset stroke (OR = 0.96, 95% CI: 0.92-1.00; p = 0.036).

**Table 1.**
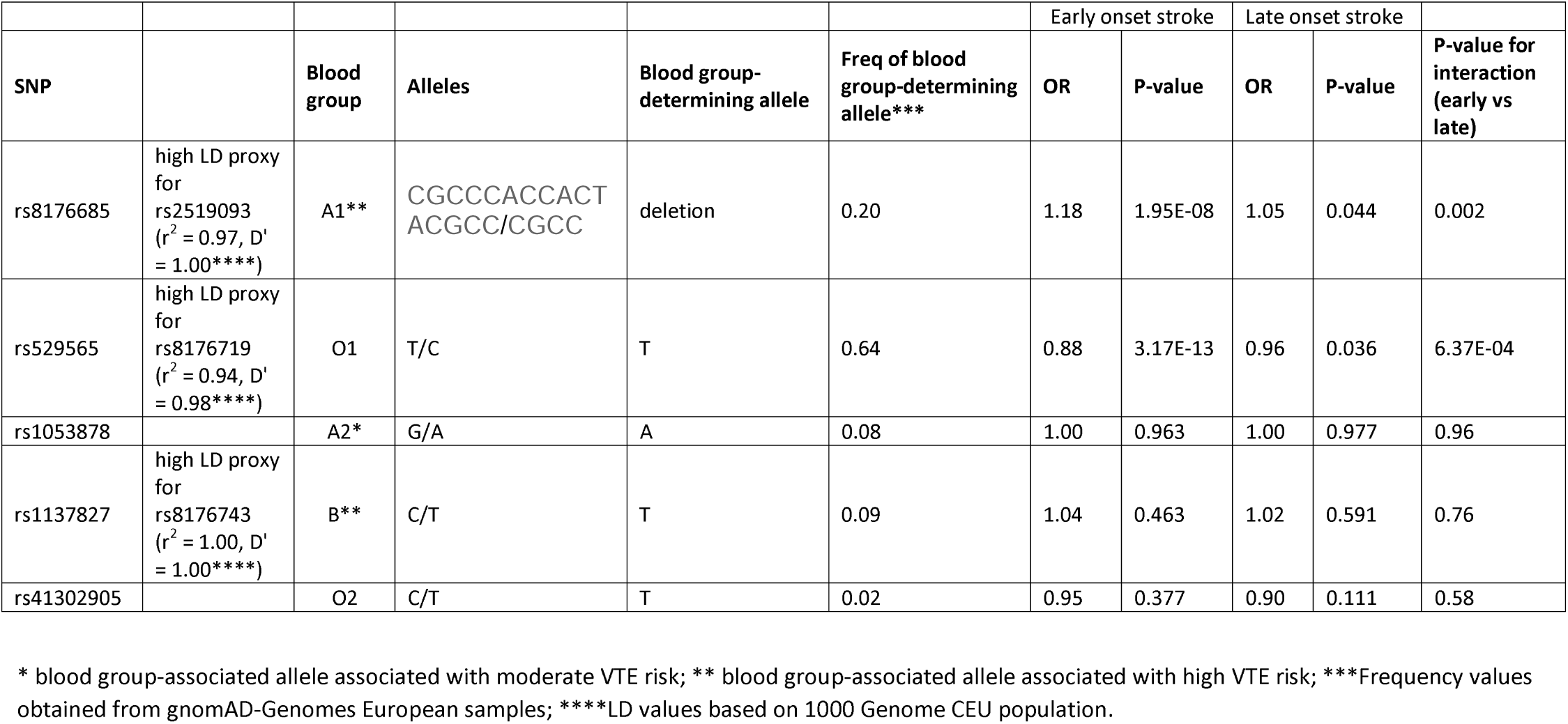
Association of the ABO blood group-defining SNPs with early and late onset stroke, EUR only

To evaluate if rs8176685 and rs529565 accounted for all of the genetic effects at the ABO locus, we performed a conditional analysis in Europeans only of the ABO locus to test for association of all SNPs at the *ABO* locus (± 50 kb from the gene) with early onset IS after including rs8176685 and rs529565 in the model as covariates. These analyses revealed 7 SNPs, falling within 4 LD groups, to be associated with all IS at a p-value < 0.01. Of these, rs5598407 (MAF = 0.045) was the most strongly associated with IS (OR = 1.21, p = 3.13 × 10^−4^). This SNP was in LD with all of the blood group-defining SNPs (r^2^ < 0.03 but D’ = 1 for all), and with rs176694 (r^2^=0.347), which is strongly associated with E-selectin levels (p < 10^−406^) in the GWAS catalogue.^27^ None of the tag SNPs for the 3 other blood group haplotypes (rs1053878-A2, rs1137827-B, and rs41302905-O2) showed evidence for association after conditioning on rs8176685 and rs529565 (p > 0.40 for all), which tag blood groups A1 and O1, respectively.

### Association of haplotype-defined ABO serologic blood group with early and late onset stroke

We assessed the relation of serologic blood groups at the *ABO* locus (i.e., A, B, AB, O blood groups) to stroke by comparing the distribution of blood types between early onset IS cases, late onset IS cases, and non-stroke controls, defining the blood groups A, B, and O genetically using a combination of SNPs ^19^. These comparisons revealed that the distribution of blood groups differed significantly between early and late onset stroke cases (p = 0.0001), between early onset stroke cases and controls (p < 0.00001), and between late onset stroke cases and controls (p = 0.0009) (Table 2). Pairwise comparisons then revealed the frequency of blood group A to be more common in early onset compared to late onset stroke (48.4% vs 45.2%; p = 0.0001) and least common in non-stroke controls (44.4%; p < 0.00001 for early onset stroke vs controls and p = 0.20 for late onset stroke vs controls). Conversely, blood group O was less common in early compared to late onset stroke cases (35.5% vs 39.1%, p < 0.0001), and most frequent in non-stroke controls (41.1%, p < 0.0001 compared to both early and late onset stroke cases). The frequency of blood group B was slightly higher in both early (11.3%) and late (11.0%) onset stroke cases compared to non-stroke controls (10.1%) (p = 0.004 and 0.012 respectively). We considered the possibility that this association could be due to a survival bias (i.e., a preferential survival of those with blood group O by assessing the association of age with blood group). Among non-stroke controls, age group (age < 60 vs ≥ 60 years) was not associated with blood group, implying that the association of early onset stroke with blood group was not driven by differential survival between subjects with different blood groups.

**Table 2.**
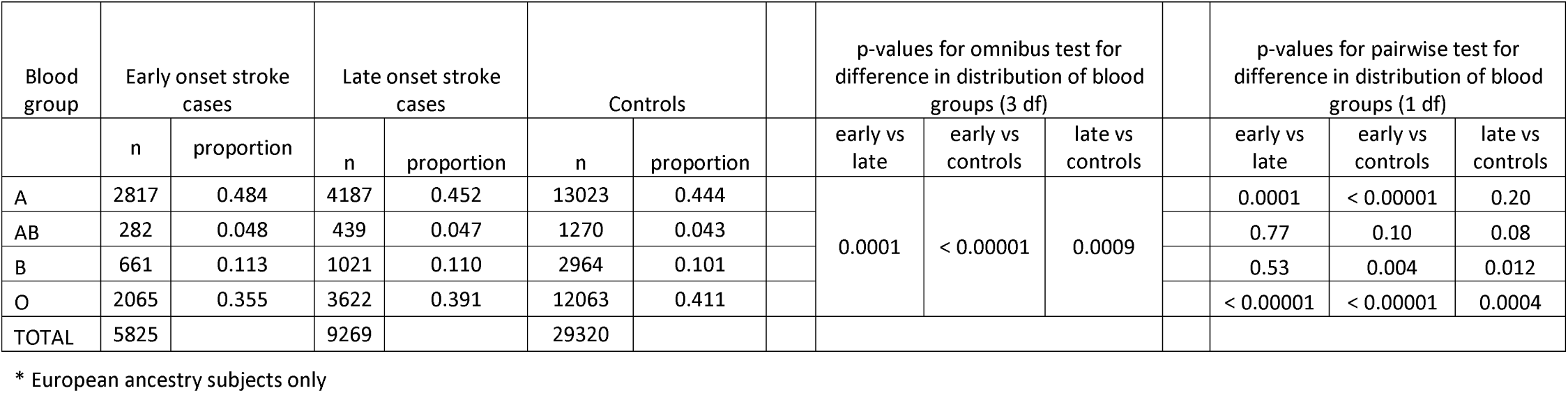
Distribution of ABO blood types in early onset stroke cases, late onset stroke cases, and in non-stroke controls*

### Associations of other established stroke loci with early onset stroke

We compared the effect sizes of 40 loci previously found to associate with IS ^1, 2, 16^ between early (age at first stroke < 60) and late (≥ age 60) onset stroke. As indicated in eTable 7, the odds ratios associated with EOS were generally consistent with those estimated for later onset stroke with two exceptions, *RGS7* rs146390073 and *TM4SF4* rs7610618, although the minor allele frequencies were relatively rare for both SNPs and there were no statistically significant differences between early and late onset IS.

### Genetically defined ABO and risk of early and late onset VTE

Because the *ABO* locus has been previously associated with VTE and other prothrombotic states ^28^, we assessed whether there was a similar graded age-at-onset association between *ABO* SNPs rs2519093 (encoding blood group A1) and rs529565 (encoding blood group O1) and VTE in the UKB. The *ABO* rs2519093-A1 allele was more strongly associated with early onset VTE (age < 60 years; n = 3,514 cases) (OR = 1.64, 95% CI: 1.54-1.74; p = 1.42 × 10^−54^) than with late onset VTE (≥ age 60 years; n = 5,043 cases) (OR = 1.34, 95% CI: 1.27-1.41; p = 2.89× 10^−29^); p-value for homogeneity of ORs = 4.01 × 10^−7^. Similarly, the *ABO* rs529565-O1 allele was more strongly associated with early onset VTE (OR = 0.66, 95% C.I.: 0.62-0.69; p = 2.95× 10^−58^) than with late onset VTE (OR = 0.77, 95% C.I.: 0.74-0.81; p = 6.63× 10^−28^); p-value for homogeneity of OR = 2.15 × 10^−6^) (details in eMethods).

### Genetic risk of VTE on risk of early and late onset stroke

We also used 2 approaches to evaluate whether genetic risk of VTE confers risk for stroke, and especially for earlier onset stroke. First, according to LDSC analysis on our current GWAS and the GWAS of VTE from INVENT, the genetic correlation between VTE and early onset stroke was 0.376 ± 0.153, p = 0.014 and between VTE and late onset stroke was 0.032 ± 0.198, p = 0.871. The genetic correlation between early and late onset stroke was 0.455 ± 0.128, p = 0.0004).

Second, genetic risk of VTE was measured as a 255-SNP-based polygenic risk score (PRS), ^23^ as described in the eMethods. A 1-SD unit increase in VTE PRS was associated with a 1.13-fold increase in risk of early onset stroke (95% C.I.: 1.10 -1.16, p < 5.21× 10^−16^) and a 1.04-fold increase in risk of late onset stroke (95% C.I.: 1.01 -1.08, p = 0.010); p-value for homogeneity of OR = 0.0002). These results were essentially unchanged when the analysis was repeated after removing 7 SNPs at the *ABO* locus from the PRS.

### Co-location of associations at the ABO locus between EOS and other thrombotic-related disorders and biomarkers

Figure 3 provides a visualization of the SNPs most strongly associated with early onset stroke and their corresponding associations with VTE and two prothrombotic biomarkers. Each panel shows the zoom plot at the *ABO* locus depicting the association of SNPs at the *ABO* locus (± 50 kb) with early onset IS. The heat map in each panel and the color coding of the variants depict the association with late onset stroke (Panel 3A), VTE (Panel 3B), plasma levels of von Willebrand factor (VWF, Panel 3C), and plasma levels of Factor VIII (F8, Panel 3D). The SNPs most strongly associated with EOS tended also to be the ones most strongly associated with VTE, VWF, and F8. This trend was less apparent for late onset stroke. Consistent with these observations, there was strong evidence using formal colocalization analyses to support the hypothesis of colocalization of at least 1 shared causal SNP between EOS and VTE, EOS and VWF, and EOS and FVIII (posterior probability supporting H4 > 99% for all pairs). Consistent with the weak and disperse set of associations with late onset stroke at this locus, there was insufficient evidence to strongly support either colocalization or absence of colocalization of shared causal SNPs between EOS and late onset stroke (posterior probability supporting H4 = 39%; posterior probability supporting H3 = 61%).

**Figure 2.**
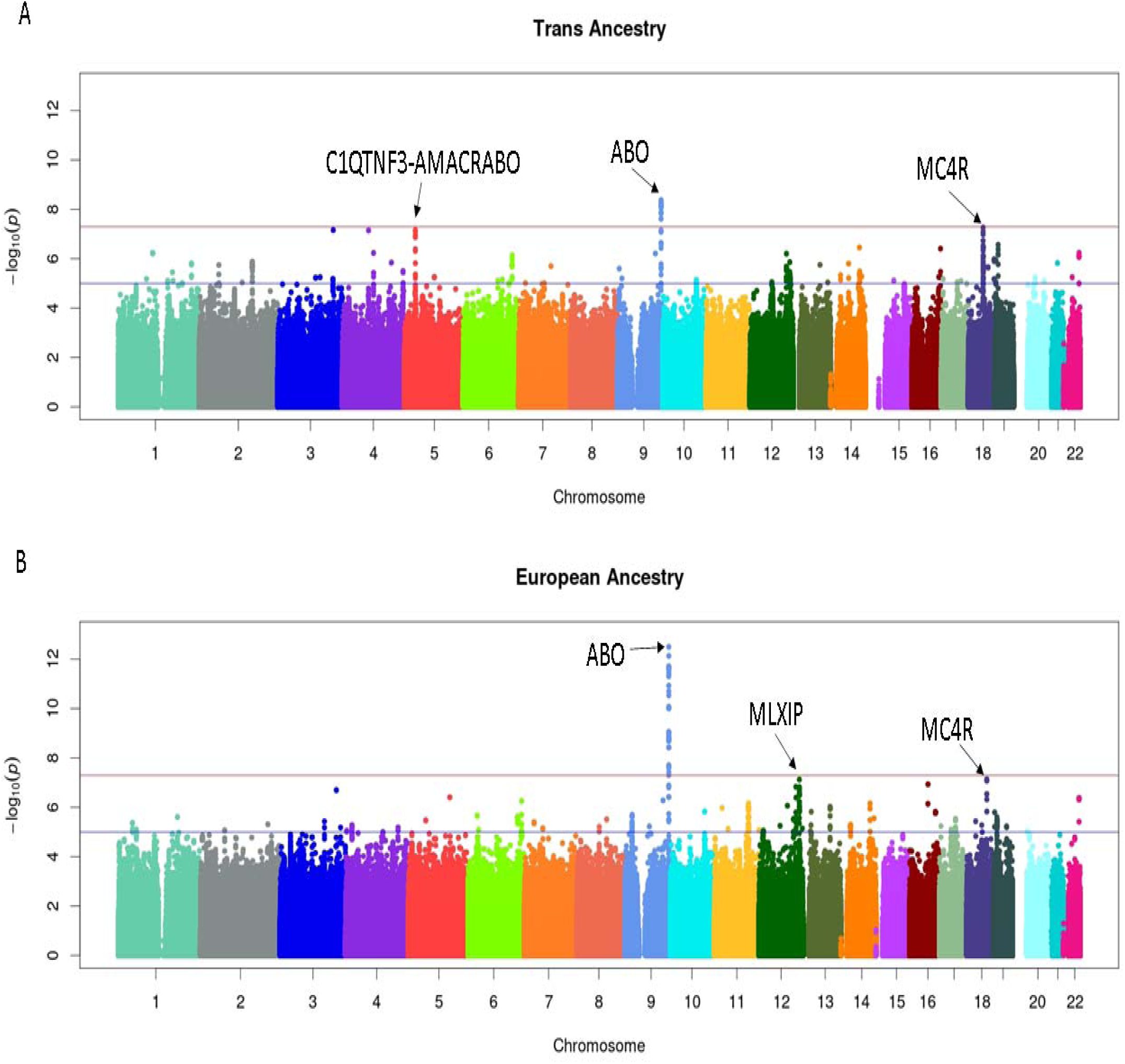
Manhattan plot of early onset stroke meta-analysis in the EOSC, random effects model, in TRANS ancestry (panel A) and European ancestry (panel B) showing association with *ABO*. The orange line marks the GWAS significant threshold of 5×10^−8^ and the black line marks the GWAS suggestive threshold of 1×10^−6^.

**Figure 3:**
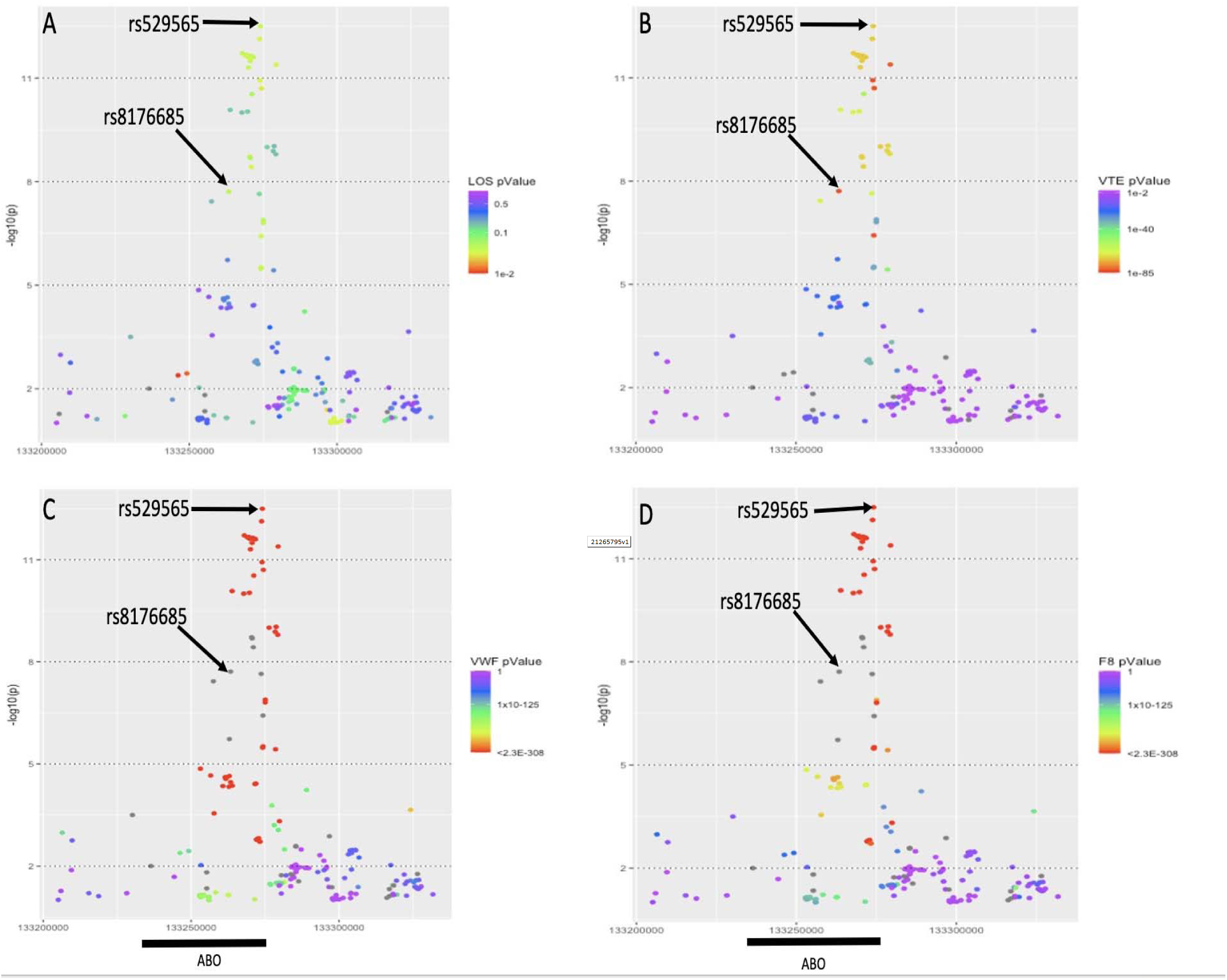
Association plots showing SNPs associated with early onset stroke at the *ABO* locus (± 50 kb) and the corresponding associations of these SNPs with late onset stroke (Panel 3A), VTE (Panel 3B), plasma levels of von Willebrand factor (Panel 3C), and plasma levels of Factor VIII (Panel 3D).

## Discussion

Our analyses revealed two variants at the *ABO* locus that were highly associated with early onset IS. These variants tag two of the ABO blood groups, blood group A1 and O1, showing a strong deleterious and protective association with IS, respectively. Non-O blood groups have been associated previously with risk of IS,^29-31^ but the novel contributions of our analysis are in showing a significantly stronger association of these blood groups with early compared to late onset stroke and in linking risk predominantly to blood group haplotype A1. In particular, our analyses suggest that the haplo-blood group A1 and O1-tagging variants rs8176685 and rs529565 are sufficient for capturing nearly all of the *ABO*-mediated genetic association with early (and perhaps late) onset stroke. Stratified analyses indicate that both SNPs are independently associated with stroke and further association analyses at the *ABO* locus that condition on the effects of these two SNPs reveal only modest additional signal at this locus.

It should be highlighted that the stronger association of *ABO* in early compared to late onset stroke is seen on the relative scale. That is, the associations observed in our study were estimated using logistic regression in case-control study designs. Since the absolute risk of stroke is smaller at younger ages, it is unclear if the larger differential effect is seen also on the additive scale. Future prospective studies that can estimate risk directly are needed to investigate the absolute risks of IS incurred by *ABO*.

The association of *ABO* in early onset stroke suggests that the associations of *ABO* with IS may be mediated via thrombotic mechanisms and not atherosclerosis. Supporting this speculation, prior studies have shown *ABO* rs687289, which is in high LD with rs529565 and tags blood group O1, to be strongly associated with plasma levels of coagulation factor VIII and its carrier protein, von Willebrand factor ^32^. These factors regulate hemostasis and thrombosis, and higher levels are associated with risk of arterial and venous thrombosis ^33^. We have further shown in our analyses of UKB data that *ABO* rs529565 is more strongly associated with early compared to late onset VTE. Genetic correlations and PRS further support that genetic risk of VTE, a well-recognized prothrombotic-related disorder, is also more strongly associated with early compared to late onset stroke.

Although our study had limited power to examine IS subtypes, it is notable that the peak *ABO* SNP in the trans-ancestry meta-analysis, rs8176685, was also significantly associated with the subtypes of large artery atherosclerosis, cardioembolism, and undetermined. This SNP was not associated with the small artery occlusion or the other determined subtypes, although the latter subtype had a very small samples size. Perhaps because of the weaker association, in MEGASTROKE, the *ABO* locus was associated only with all IS but not with any subtype.

Differential associations of the *ABO* locus with myocardial infarction and coronary artery calcification further support a prothrombotic mechanism. The *ABO* locus has been associated with susceptibility to myocardial infarction (ICD-10 code 411.2) in CARDIoGRAM ^34^ and in the UKB (p = 2.1E-5; from https://pheweb.org/UKB-TOPMed/variant/9:133274084-C-T, Accessed 4/5/2021). However, there is no evidence for a strong association of this locus with coronary artery calcification, a marker for subclinical atherosclerosis ^35, 36^, implying that this association may be mediated by a mechanism other than atherosclerosis (e.g., hemostasis). Taken together, our analyses with these additional observations support an increased genetic role of prothrombotic mechanisms in early compared to later onset stroke. This speculation is further supported by our colocalization analyses supporting the presence of a shared causal variant at the ABO locus between EOS and VTE, EOS and VWF, and EOS and FVIII. Evidence supporting a shared causal variant at the ABO locus between EOS and late onset stroke was equivocal, perhaps because of the relatively low strength of association between late onset stroke and the *ABO* locus and consequently low power of the colocalization test.

This leads to the question, what are the clinical implications of an enrichment of prothrombotic mechanisms in early onset stroke? Clinical translation will require a better understanding of the prothrombotic mechanisms in early-onset stroke and, likely, a personalized secondary prevention strategy. The effect sizes of the stroke-associated common variants at the *ABO* locus are too small per se to have immediate clinical implications, but gene-gene and gene-environment interaction deserve future study^37^. One path to translation would be to identify gene-drug interactions (e.g. oral contraceptives and genetic risk for thrombosis) and determine whether the joint effect has implications for primary prevention. Additional research implications are that rare variant studies should target prothrombotic and related pathways, which could identify variants of larger effect size.

In addition to *ABO*, we detected genome-wide evidence for association of early onset stroke with *MLXIP* rs377424471. This locus has not previously been associated with stroke to our knowledge, nor did rs377424471 show any evidence for association with late onset stroke in SiGN. *MLXIP* does, however, play a role in glucose-responsive gene regulation, making it a plausible candidate gene for stroke.^38^ The association of this SNP with early onset stroke requires future replication in an independent early onset stroke cohort.

Our study is not without limitations. Although 35% of subjects in the EOSC are of non-European ancestry, the diversity of the current EOSC cohort is still somewhat limited, reducing power to detect variants whose frequencies might be high in non-European populations yet low in Europeans. A second major limitation is that the sample size for all IS is still small by GWAS standards and power to detect subtype-specific variants is even more limited.

In summary, our genome-wide analysis indicates a stronger association of *ABO* risk variants tagging blood groups A1 and O1 with earlier compared to later onset stroke, and stronger associations of the same ABO variants with early compared to late onset VTE, another prothrombotic condition. Similarly, we observed genetic risk for VTE to be more strongly correlated with early compared to late onset stroke. Our findings are consistent with an increased role for prothrombotic mechanisms in early compared to later onset stroke.

## Study funding

This study was supported by the National Institutes of Health grants R01 NS105150, R01 NS100178 and R01NS114045 and Department of Veterans Affairs grant BX004672-01A1. The project utilized the resources and facilities at the VA Maryland Health Care System, Baltimore, Maryland.

The contents do not represent the views of the U.S. Department of Veterans Affairs or the United States Government.

Tom Jaworek was supported by a T32 AG000262 Epidemiology of Aging grant. Huichun Xu was supported by AHA Career Development Award (19CDA34760258).

Please see eAppendix 1 for the funding and acknowledgements for each contributing study.

## Supporting information

Table of Contents

