## Supplementary material for "Genetic Contributions to Early and Late Onset Ischemic Stroke": Figure 1

**Genome-Wide Association Analysis of Young-Onset Stroke Identifies a Locus on Chromosome 10q25 Near *HABP2*. *Cheng et al., Stroke 2016***

eFigure 1. Association of HABP2 with early onset stroke in the EOSC

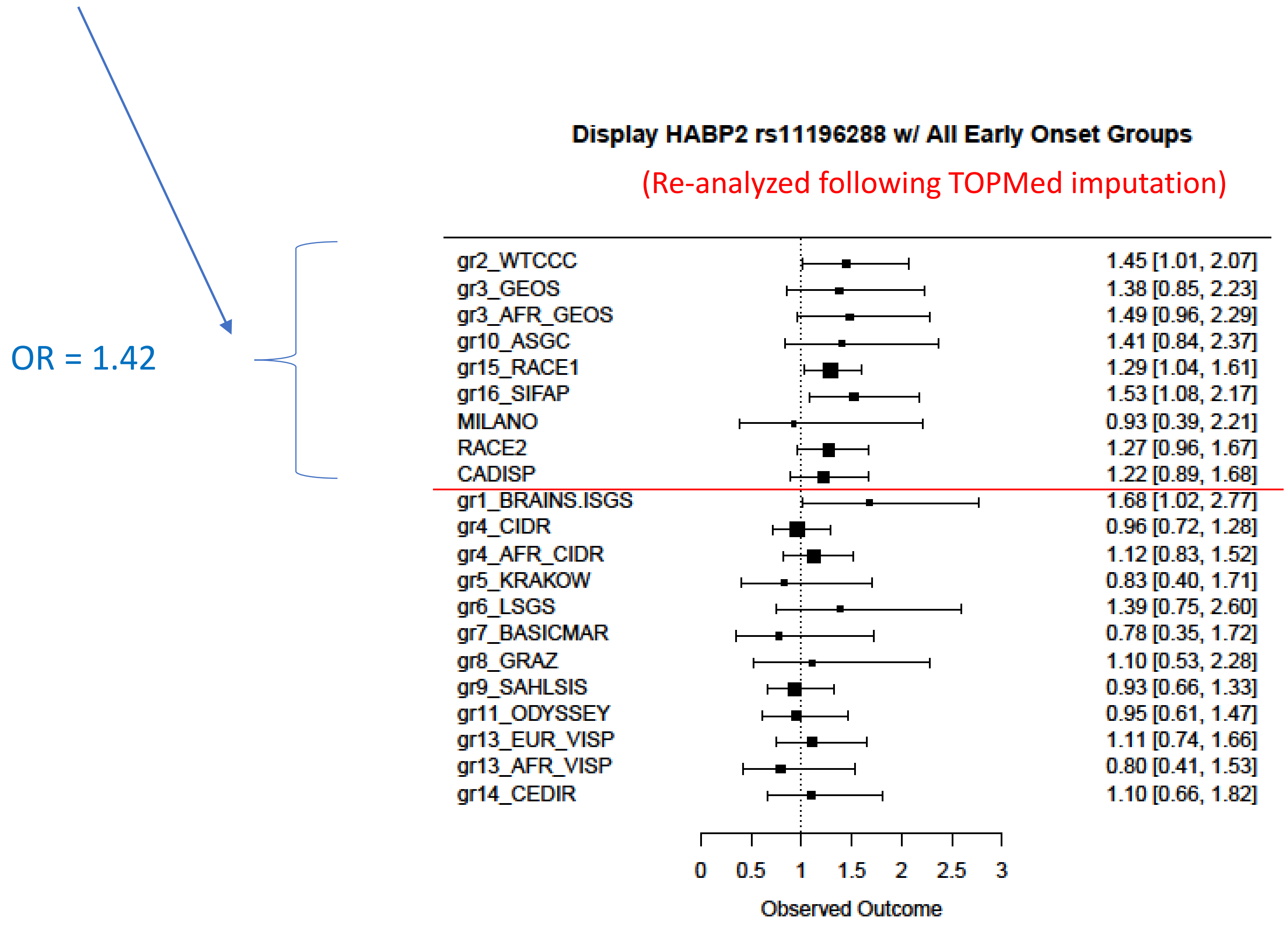
