## Supplementary material for "Genetic Contributions to Early and Late Onset Ischemic Stroke": Figure 2a

eFigure 2a. Association of *ABO* SNP rs8176685 (blood group A1) with early and late onset ischemic stroke (TRANS analysis)

| | N Cases | OR | L95 | U95 | p-value | p-value (homogeneity) |
|---|---|---|---|---|---|---|
| **All stroke** | 16248 | 1.167 | 1.114 | 1.223 | 8.25E-11 | 0.008 |
| | 9832 | 1.052 | 0.990 | 1.118 | 0.103 | |
| **SAO** | 3190 | 1.104 | 0.945 | 1.288 | 0.212 | 0.764 |
| | 1777 | 1.071 | 0.955 | 1.201 | 0.238 | |
| **LAA** | 1900 | 1.170 | 1.032 | 1.326 | 0.014 | 0.196 |
| | 1747 | 1.054 | 0.957 | 1.161 | 0.285 | |
| **CE** | 2038 | 1.243 | 1.095 | 1.410 | 7.45E-04 | 0.126 |
| | 2665 | 1.107 | 1.026 | 1.195 | 0.008 | |
| **Other** | 886 | 1.101 | 0.923 | 1.314 | 0.285 | |
| | 71 | NA | NA | NA | NA | |
| **Undetermined** | 5149 | 1.201 | 1.105 | 1.306 | 1.69E-05 | 0.011 |
| | 2182 | 0.969 | 0.840 | 1.118 | 0.668 | |

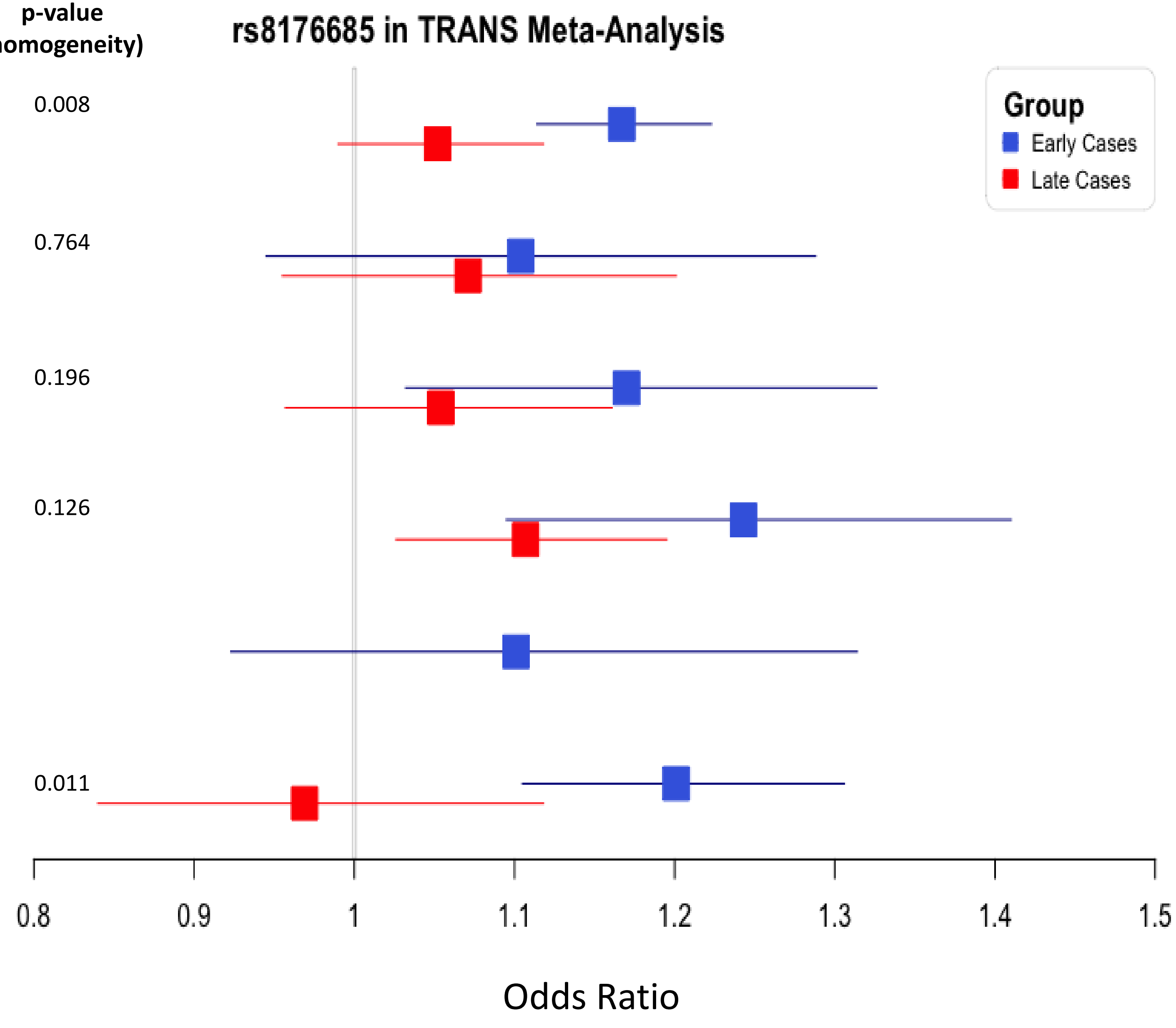
