## Supplementary material for "Genetic Contributions to Early and Late Onset Ischemic Stroke": Figure 2b

eFigure 2b. Association of *ABO* SNP rs529565 (blood group O1) with early and late onset ischemic stroke (EUR analysis)

| | N Cases | OR | L95 | U95 | p-value | p-value (homogeneity) |
|---|---|---|---|---|---|---|
| **All stroke** | 11311 | 0.877 | 0.847 | 0.925 | 3.17E-13 | 5.34E-04 |
| | 9272 | 0.961 | 0.909 | 0.998 | 0.036 | |
| **SAO** | 1346 | 1.035 | 0.929 | 1.153 | 0.535 | 0.297 |
| | 1627 | 0.964 | 0.891 | 1.043 | 0.363 | |
| **LAA** | 791 | 0.784 | 0.694 | 0.887 | 1.03E-04 | 4.39E-02 |
| | 1681 | 0.911 | 0.842 | 0.985 | 0.019 | |
| **CE** | 953 | 0.814 | 0.727 | 0.912 | 3.78E-04 | 4.21E-02 |
| | 2580 | 0.932 | 0.873 | 0.994 | 0.033 | |
| **Other** | 376 | 0.950 | 0.814 | 1.107 | 0.506 | 0.825 |
| | 63 | 0.932 | 0.873 | 0.994 | 0.033 | |
| **Undetermined** | 1502 | 0.867 | 0.805 | 0.935 | 1.92E-04 | 0.002 |
| | 2058 | 1.021 | 0.951 | 1.096 | 0.569 | |

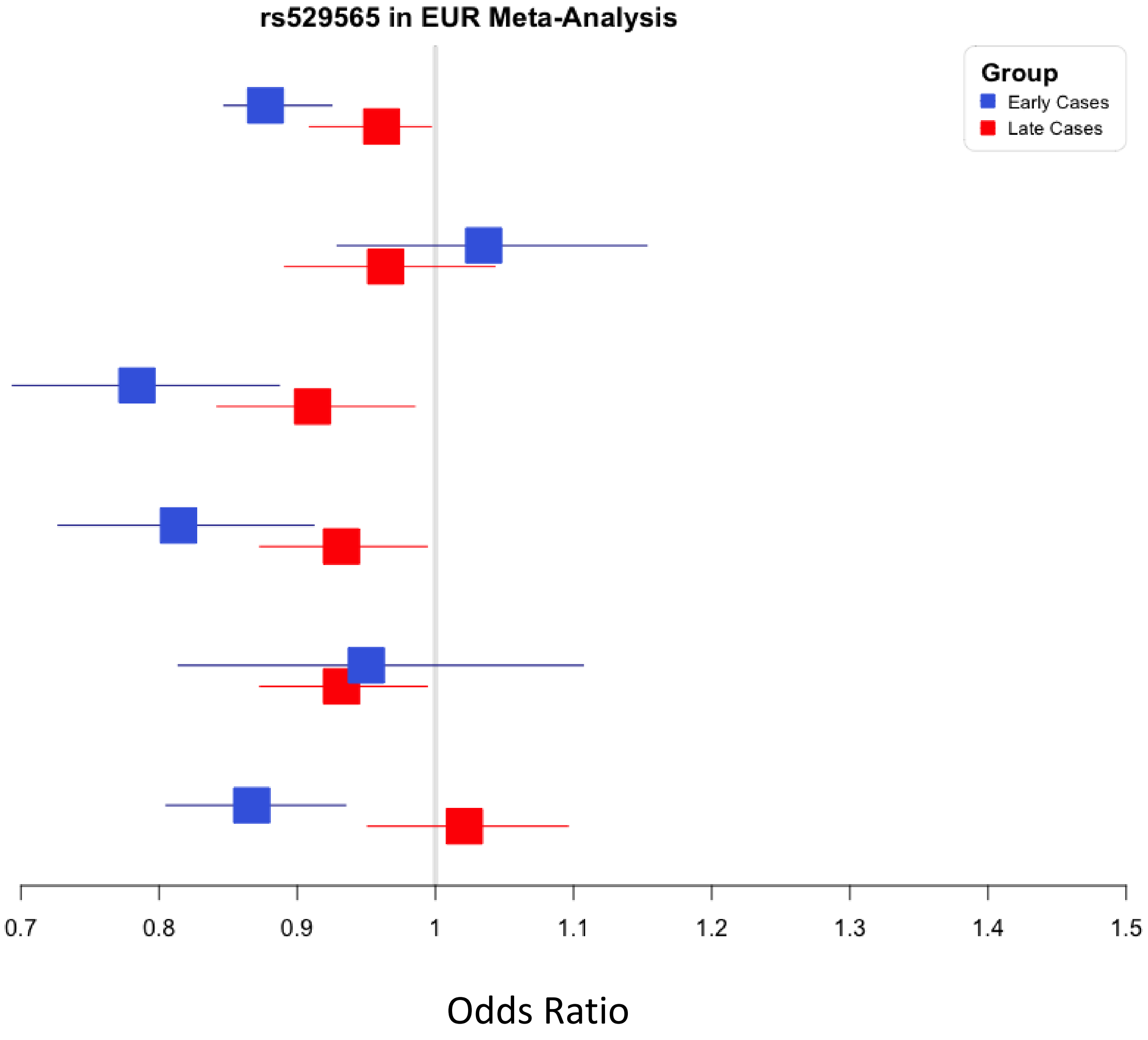
