## Supplementary material for "Genetic Contributions to Early and Late Onset Ischemic Stroke": Figure 3

### eFigure 3. Locuszoom plots of 2 SNPs associated with AIS in EUR

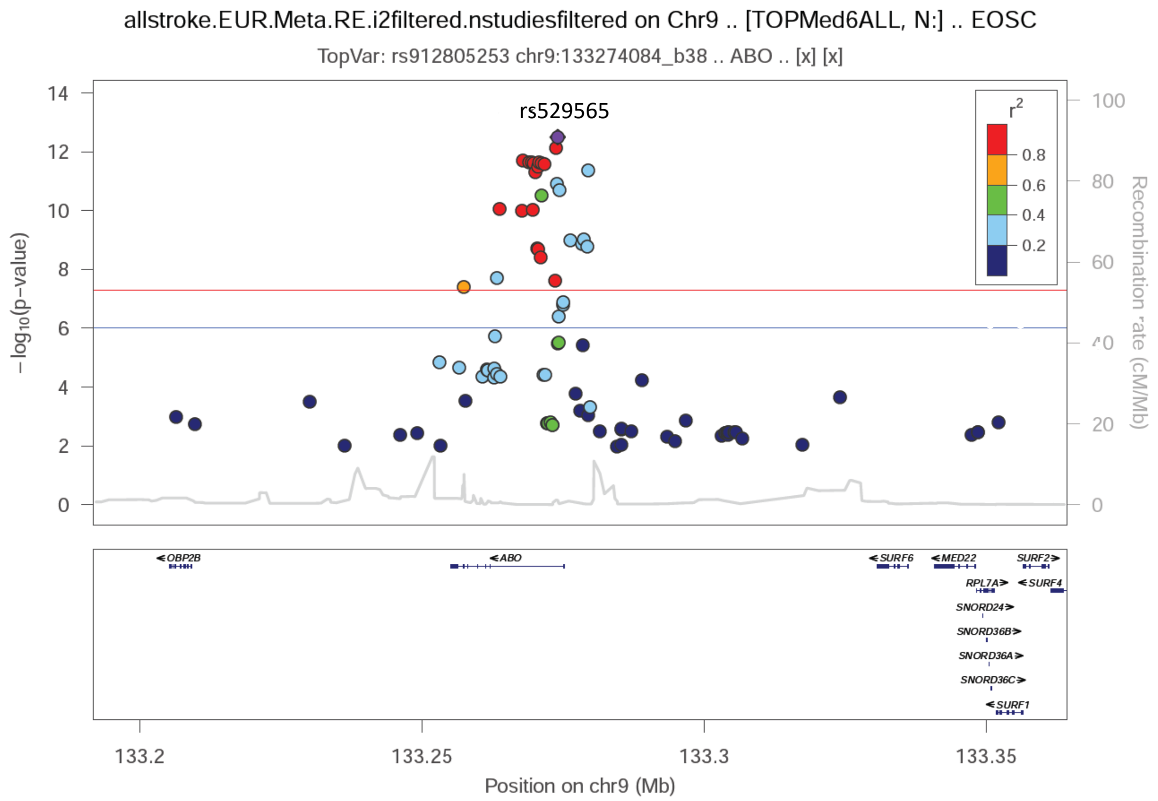


rs529565 is top SNP from EUR metaanalysis

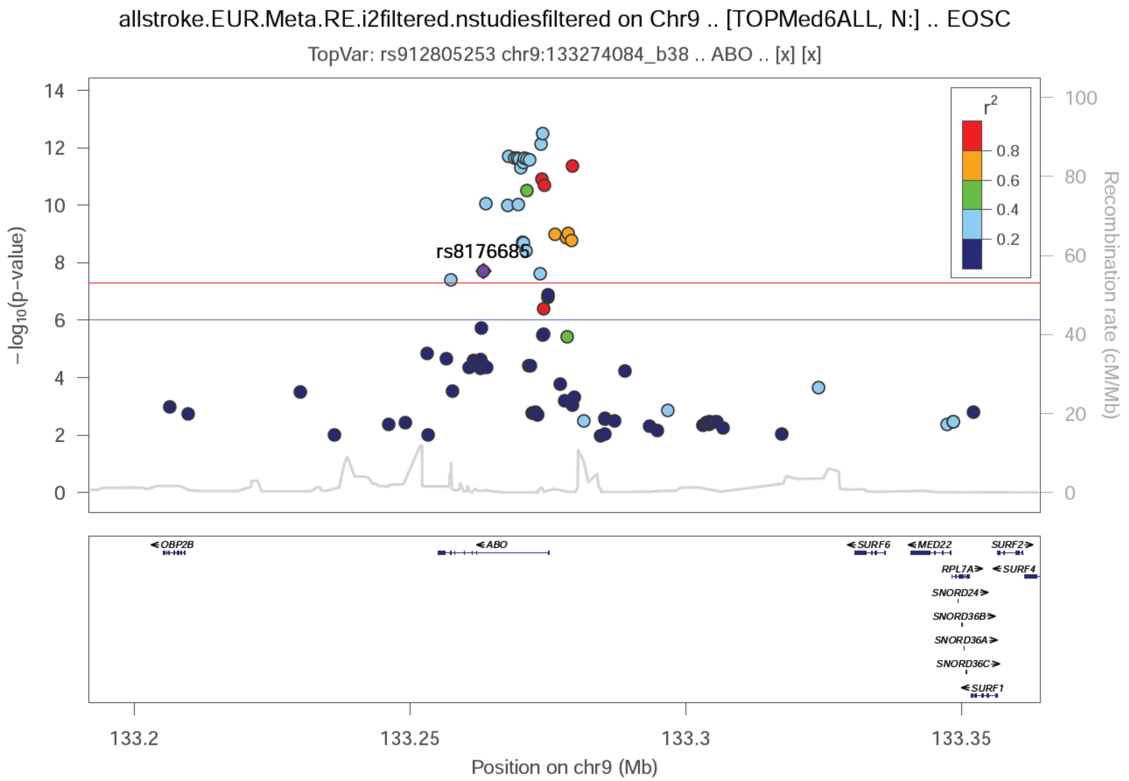


rs8176685 is top SNP from TRANS metaanalysis
