## Supplemental Methods for "Genetic Contributions to Early and Late Onset Ischemic Stroke"

**eMethods**

Table of Contents

1. Cohort Descriptions	1
   1.1. Case cohorts	1
   1.2. Control-only cohorts	21
2. Genome-wide association analysis	25
3. Conditional analysis at the ABO locus	25
4. ABO blood group-determining SNPs, serologic ABO blood groups, and stroke	26
   4.1. Association of ABO haplotype-defining SNPs with stroke	26
   4.2. Distribution of ABO blood group among early onset stroke, late onset stroke, and controls	26
5. Association of ABO SNPs with early and late onset stroke in UKB	27
6. Association of ABO SNPs with early and late onset VTE in UKB	27
7. Association of VTE PRS with early and late onset stroke	28
8. Co-location of associations at the ABO locus between EOS and other thrombotic-related disorders and biomarkers	28
9. References	30
1. **Cohort Descriptions**

**Australian Stroke Genetics Collaborative (ASGC)**

ASGC stroke cases comprised European-ancestry stroke patients admitted to four clinical centres across Australia (The Neurosciences Department at Gosford Hospital, Gosford, New South Wales (NSW); the Neurology Department at John Hunter Hospital, Newcastle, NSW; The Queen Elizabeth Hospital, Adelaide; and the Royal Perth Hospital, Perth) between 2003 and 2008. Stroke was defined by WHO criteria as a sudden focal neurologic deficit of vascular origin, lasting more than 24 hours and confirmed by imaging such as computerized tomography (CT) and/or magnetic resonance imaging (MRI) brain scan. Other investigative tests such as electrocardiogram, carotid doppler and transoesophageal echocardiogram were conducted to define IS mechanism as clinically appropriate. Cases were excluded from participation if aged <18 years, diagnosed with haemorrhagic stroke or transient ischemic attack rather than IS, or were unable to undergo baseline brain imaging. IS subtypes were assigned using TOAST criteria, based on clinical, imaging and risk factor data. Consenting participants completed five detailed self-report questionnaires and attended the HCS data collection centre, at which time a series of clinical measures were obtained. All study participants gave informed consent for participation in genetic studies. Approval for the individual studies was obtained from relevant institutional ethics committees.

ASGC controls were participants in the Hunter Community Study (HCS), a population-based cohort of individuals aged 55-85 years, predominantly of European Caucasian ancestry and residing in the Hunter Region, NSW, Australia. Briefly, participants were randomly selected from the NSW State electoral roll and contacted by mail between 2004 and 2007.

**BASEede datos de ICts del hospital del MAR (BASICMAR)**

BASICMAR is an ongoing prospective study of all acute strokes assessed since 2005 at the IMIMHospital Universitari del Mar (Barcelona, Spain). It includes both first-ever and recurrent strokes. There were no exclusion criteria regarding age or race-ethnicity of the individuals. All patients had an electrocardiogram (ECG), a blood analysis, and neuroimaging at the acute stage. Additional diagnostic procedures were performed when clinically indicated. A follow-up of three months after stroke was completed for all survivors. Ischemic stroke etiologic subtypes were classified according to TOAST criteria. For this study, only individuals of European origin with ischemic stroke were selected from BASICMAR, with eligible events defined as a clinical syndrome of any duration associated with a radiographically proven acute infarct, without radiographic evidence of a demyelinating or neoplastic disease or other structural disease including primary intracerebral hemorrhage.

**Biobank Japan**

BioBank Japan Project was started in 2003 and collected DNA and clinical information from a total of 200,000 patients suffering from at least one of 47 common diseases, including ischemic stroke over age 40. Ischemic stroke was diagnosed by physicians at the 66 collaborating hospitals and its subtypes were determined by medical record review according to the TOAST criteria.

For controls, we used genotyping information of 27,294 individuals aged over 40 years from three population-based controls: Japan Multi-Institutional Collaborative Cohort Study(J-MICC), Japan Public Health Center-based Prospective Study (JPHC), and the Tohoku Medical Megabank Project (TMM).

**Biorepository of DNA in Stroke (BRAINS)**

BRAINS is a hospital-based study. Cases participating in the current study were of European descent and recruited within the United Kingdom between September 25, 2009 and August 4, 2011. Extensive phenotype information is collected including subtype of stroke, past and family cardiovascular history, blood pressure data, MRI or CT brain imaging, carotid anatomy, and blood tests (including cholesterol). All hospital admitted participants over the age of 18 years with first-ever or recurrent stroke that provided informed consent (or caregivers on their behalf) were recruited. Participants must have image-positive lesions. Exclusion criteria are mainly for those that were brain image-negative, even if the clinical presentation is that of stroke. There are no eligibility criteria based on stroke severity or participation in a treatment trial. Inability to obtain consent results in mandatory exclusion.

Controls were European-Ancestry, stroke-free participants from the shared WTCCC controls, a prospectively collected cohort of individuals born in 1958 (1958 Birth Cohort).

**Cervical Artery Dissections and Ischemic Stroke Patients (CADISP)**

The CADISP Study aims primarily at identifying genetic and environmental risk factors for cervical artery dissection (CeAD), a major cause of ischemic stroke in young adults. We included 942 CeAD patients in the CADISP study in 2004-2009 (CADISP-1; 170 Finns and 772 non-Finnish Europeans). Additional 451 CeAD patients of European origin were recruited in 2008-2010, exclusively for the CADISP-genetics project, in some CADISP centers and additional European and US centers (CADISP2). To assess the specificity of risk factors for CeAD, we also recruited 583 patients with an IS attributable to other causes (non-CeAD IS, 162 Finns and 421 non-Finnish Europeans from Belgium, France, Germany, Italy, and Switzerland), frequency-matched on age, sex and geographical origin with CADISP-1 CeAD patients. Of these, a total of 555 non-CeAD IS patients aged < 60 years, who were successfully genotyped and met genotyping quality control criteria, were available for the present analysis. The abstracted hospital records of cases were reviewed and adjudicated for IS subtype by a neurologist in each participating center. Each item required for the subtype classification was also recorded in a standardized fashion. Based on this, IS subtypes were then centrally readjudicated by a panel of neurologists, in agreement with the TOAST system, using the more detailed subtype description from Ay et al. ^1^ DNA samples were genotyped on an Illumina Human610-Quad or Human660W-Quad BeadChip® at the Centre National de Génotypage [CNG], Evry, France.

The majority of controls (N=9,046, of which 74 Finns and 8,972 non-Finnish Europeans) were selected from an anonymized control genotype database at the CNG, in order to match cases for ethnic background, based on principal component analysis. European reference samples from the genotype repository at the CNG were also analyzed simultaneously to provide improved geographical resolution. Additional Finnish controls were recruited within the CADISP study, both from the general population and among spouses and unrelated friends of CADISP patients, within the Helsinki area. A total of 234 individuals were eligible for genotyping at the CNG. Of these, 213 individuals who were genotyped successfully and met quality control criteria were available for the present analysis. Individuals were excluded if they were unexpected duplicates, gender discrepancy and unexpected relatedness. After quality control, we performed genotype imputation to the nonmonomorphic SNPs described in the 1000 genomes phase I v3 reference panel.

**Edinburgh Stroke Study**

Between 2002 and 2005, consecutive consenting patients with stroke who were admitted to or seen as outpatients at the Western General Hospital, Edinburgh were prospectively recruited from stroke centers in Edinburgh, Scotland, U.K. There were no exclusion criteria for cases based on age, stroke severity, or inclusion in other clinical research studies. Cases in this study were of European origin, with a clinically evident stroke, demonstrated by brain imaging (CT or MRI) to be ischemic. An experienced stroke physician assessed each participant as soon as possible after stroke onset, prospectively recording demographic and clinical details, including vascular risk factors and results of brain imaging and other investigations. Ischemic subtypes were assigned according to the TOAST criteria and, subsequently, using CCS, specifically for the purposes of the SIGN study. ESS cases were collected as part of the WTCCC2 effort. All WTCCC2 cases were genotyped as part of the WTCCC2 Ischemic Stroke study using the Illumina Human660W-Quad array. Quality control procedures in the WTCCC2 excluded SNPs not genotyped on all case and control collections and SNPs with Fisher information measure < 0.98, genotype call rate < 0.95, MAF < 0.0 or Hardy-Weinberg P-value < 1 x 10-20 in either the case or control collections. Samples were excluded if identified as outliers on call rate, heterozygosity, ancestry and average probe intensity based on a Bayesian clustering algorithm. Samples were also removed if they exhibited discrepancies between inferred and recorded sex or were shown to have cryptic relatedness with other WTCCC2 samples (pairwise identity-by-descent > 0.05).

Controls from the UK samples were drawn from shared WTCCC controls obtained from the 1958 Birth Cohort. This is a prospectively collected cohort of individuals born in 1958 (<http://www.b58cgene.sgul.ac.uk/>) and ascertained as part of the national child development study (http://www.cls.ioe.ac.uk/studies.asp?section=000100020003).

**EPIC-CVD**

EPIC is a multi-centre prospective cohort study of 519,978 participants (366,521 women and 153,457 men, mostly aged 35–70 years) recruited between 1992 and 2000 in 23 centres located in 10 European countries. Participants were invited mainly from population-based registers (Denmark, Germany, certain Italian centres, the Netherlands, Norway, Sweden, UK. Other sampling frameworks included: blood donors (Spain and Turin and Ragusa in Italy); screening clinic attendees (Florence in Italy and Utrecht in the Netherlands); people in health insurance programmes (France); and health-conscious individuals (Oxford, UK). About 97% of the participants were of white European ancestry. Prevalent CVD was ascertained through self-reported history of MI or angina, or registry-ascertained CVD event prior to baseline. Stroke ascertainment in EPIC: Centres were asked to ascertain suspected stroke cases from registries, hospital records or self-report (i.e. follow-up questionnaires). Stroke events were defined by ICD10 codes as follows: Ischemic I63, Haemorrhagic I61, SAH I60, Unclassified I64, Other CRBV I62, I65- I69, F01. Incident stroke cases have been defined as fatal and non-fatal. All centres have recorded cause-specific mortality through mortality registries and/or active follow-up and have ascertained and validated incident fatal and non-fatal stroke through a combination of methods. Ascertained non-fatal stroke events were validated by clinical symptoms and imaging evidence (CT/MRI) or confirmed through hospital/GP records (with assessment of notes) or confirmed through hospital records (without assessment of notes). Individuals were excluded if they had clinical symptoms, but no validation was possible e.g. there was no imaging evidence, nor GP/primary care records (without assessment of notes) or registry information. Fatal stroke events were validated either by autopsy or hospital records and death certificate or by death certificate if they died in hospital. Individuals where validation was not possible were excluded. Participants with a history of stroke or MI at baseline were excluded. No further stroke subtyping was performed.

EPIC-CVD employs a nested case-cohort design, analogous to the EPIC-InterAct study for type-2 diabetes which established a common set of referents through selection of a random sample of the entire cohort (“sub cohort”).

**Follow-Up of Transient ischemic attack and stroke patients and Unelucidated Risk Factor Evaluation study (FUTURE)**

The FUTURE study is a prospective cohort study on risk factors and prognosis of young ischemic stroke and hemorrhagic stroke among 1,006 patients, aged 18-50 years, included in the study database between 1-1-1980 and 1-11-2010. Follow-up visits at the research centre occurred from the end of 2009 until the end of 2011. Control subjects were recruited among the patients’ spouses, relatives or social environment. Information on mortality and incident vascular events will be retrieved via structured questionnaires. In addition, participants were invited to the research centre to undergo an extensive sub-study including MRI. Inclusion criteria for this consortium was ischemic stroke of presumed arterial origin. Exclusion criteria include traumatic hemorrhagic stroke, intracerebral hemorrhage in known cerebral metastasis or primary brain tumor, ischemic/hemorrhagic stroke due to cerebral venous sinus thrombosis, intracerebral hemorrhage due to ruptured cerebral aneurysm, any subarachnoid hemorrhage, and retinal infarct. Controls have to be at least 18 years old without a history of any TIA or stroke before the age of 50 at the moment of inclusion.

**Mass General/GASROS**

MGH-GASROS enrolled ischemic stroke subjects as part of a single-center prospective cohort study of consecutive patients with ischemic stroke aged ≥18 years admitted to the Massachusetts General Hospital Stroke Unit (Boston, MA, U.S.A.) between 2003 and 2011 after presenting to the emergency department within 24 hours of symptom onset. Ischemic stroke was defined as a clinical syndrome of any duration associated with a radiographically proven acute infarct consistent with a vascular pattern of involvement and without radiographic evidence of a demyelinating or neoplastic disease or other structural disease, including vasculitis, subacute bacterial endocarditis, vasospasm due to subarachnoid hemorrhage or cocaine abuse, or primary intracerebral hemorrhage. Diagnosis of acute cerebral ischemia was confirmed for all subjects in the present study by admission diffusion weighted imaging (DWI) completed within 48 hours after symptom onset. Vascular and critical care neurologists subtyped ischemic strokes by systematic medical record review using the TOAST and CCS systems. Controls were matched to cases on the basis of age, sex and race/ethnicity and drawn from stroke-free individuals who received care at primary care practices within Massachusetts General Hospital.

**Greater Cincinnati/Northern Kentucky Stroke Study (GCNKSS)**

The GCNKSS is a population-based epidemiologic study of stroke in blacks and whites that is designed to measure temporal trends and racial differences in incidence of stroke. The catchment area includes two southwestern Ohio, U.S.A., counties (Hamilton, which includes the city of Cincinnati, and Clermont to the east) and three Northern Kentucky, U.S.A., counties (Boone, Kenton, and Campbell) to the south of Cincinnati across the Ohio River. As part of the GCNKSS, for calendar years 1999 and 2005, prospective cohorts of first-ever and recurrent ischemic stroke cases were assembled using “hot pursuit” methodology at all local hospitals in the region (18 in 1999, and 17 in 2005), except for one hospital that is solely devoted to treating pediatric cases. Subjects with all degrees of severity of stroke were eligible, and no particular racial group was intentionally oversampled (about 80% were white participants and 20% black). Study research nurses prospectively screened inpatient admission and emergency department logs to identify acute ischemic stroke patients. After consent was granted from the patient or legally authorized representative, a study nurse performed an extensive interview, and a blood sample was obtained for genetic analysis. In addition, a study nurse abstracted information about the individual, the subject’s medical history, the stroke event, and imaging studies from the hospital chart. A study physician reviewed every abstract, along with the imaging studies, to verify that an acute stroke had occurred, and to classify the event according to TOAST and CCS criteria.

**Geisinger**

The study cohort was made up of participants of the Geisinger’s MyCode Community Health Initiative consisting of 946 patients with acute ischemic stroke with validated European ancestry. The informed consent was obtained for all MyCode patients. This study was approved by the Geisinger Institutional Review Board. Patient characteristics, clinical Variables, and outcome Measures were based on the neurological examination and corresponding neuroimaging. Controls were non-stroke patients with validated European ancestry.

**Genetics of Early Onset Stroke (GEOS) Study**

The GEOS study is a population-based case-control study designed to identify the genetic determinants of early-onset ischemic stroke and to characterize interactions of stroke-associated genes with environmental risk factors. Cases with a first-ever ischemic stroke were identified by discharge surveillance from one of 59 hospitals in the U.S. greater Baltimore-Washington area and by direct referral from regional neurologists.

The abstracted hospital records of potential cases were reviewed and adjudicated for ischemic stroke, ischemic stroke subtype, and modified Rankin Scale ^2^ at discharge by a pair of vascular neurologists according to previously published procedures ^3^ ^4^ with disagreements resolved by a third vascular neurologist. Stroke was defined according to the criteria of the WHO ^5^ and ischemic stroke was defined based on the criteria of the National Institute of Neurological Disorders and Stroke Data Bank.^6^ Cases had a head CT and/or brain MRI that was consistent with cerebral infarction. Visualization of the infarct was not required, only that no alternative etiology was identified. The ischemic stroke subtype classification system retains information on all probable and possible causes and is reducible to the more widely used TOAST1 system that assigns each case to a single category. Cases were subsequently subtyped using the CCS.

Ischemic strokes with the following characteristics were excluded from participation: stroke occurring as an immediate consequence of trauma, stroke within 48 hours after a hospital procedure, stroke within 60 days after the onset of a non-traumatic subarachnoid hemorrhage, and cerebral venous thrombosis. Additional exclusions for genetic analyses modified from(10) were as follows: known single-gene or mitochondrial disorders recognized by a distinctive phenotype (e.g. cerebral autosomal dominant arteriopathy with subcortical infarcts and leukoencephalopathy (CADASIL), mitochondrial encephalopathy with lactic acidosis and stroke-like episodes (MELAS), homocystinuria, Fabry disease, or sickle cell anemia); mechanical aortic or mitral valve at the time of index stroke; untreated or actively treated bacterial endocarditis at the time of the index stroke; neurosyphilis or other central nervous system infections; neurosarcoidosis; severe sepsis with hypotension at the time of the index stroke; cerebral vasculitis by angiogram and clinical criteria; post-radiation arteriopathy; left atrial myxoma; major congenital heart disease; and cocaine use in the 48 hours prior to stroke onset. There were no exclusions based on race or ethnicity, stroke severity, or participation in clinical trial research.

Control participants without a history of stroke were identified by random-digit dialing. Controls were balanced to cases by age and region of residence in each study and were additionally balanced for ethnicity in SPYW-2 and SPYM.

**GRAZ**

Between 1994 and 2003, subjects with first-ever and recurrent ischemic strokes admitted to the stroke unit of the Department of Neurology, Medical University of Graz (Graz, Austria) were included. All race-ethnic groups were eligible and there was no intentional oversampling. All age groups were allowed, though only subjects above the age of 18 were admitted to our department. Ischemic stroke was defined as an episode of focal neurological deficits with acute onset and lasting > 24 hours. There were no selection criteria based on stroke severity. Those individuals in treatment trials were excluded. 685 subjects were eligible to participate in this study (278 women, 407 men). All cases were Caucasian. Mean age was 68.9 ± 13.8 years with an age range from 19 – 101 years. In addition to a standardized protocol including a laboratory examination and carotid ultrasound or magnetic resonance angiography and ECG, 304 subjects underwent neuroimaging by CT and 381 by MRI. More extensive cardiac examination, including transesophageal echocardiography or transthoracic echocardiography and Holter, was performed in subjects with suspected cardiac embolism. Stroke subtypes were assessed according to modified TOAST criteria and were conducted by trained stroke neurologists. Controls were from the Austrian Stroke Prevention Study.

**Helsinki Ischemic Stroke Genetics Study**

Helsinki Ischemic Stroke Genetics Study was designed for investigating genetic factors underlying ischemic stroke in the Finnish population and in the long-term to be incorporated to multicenter multinational similar datasets. Ischemic stroke cases were recruited from 2012 to 2017 from the Helsinki University Hospital, Department of Neurology, Helsinki, Finland, which is the only neurological emergency unit for a population of 1.7 million inhabitants. 1848 patients with positive neuroimaging findings for a new-onset brain infarction were included. Furthermore, we enrolled young patients aged 18 to 55 years with MRI-positive acute ischemic stroke from 2008 to 2010, and of these 72 patients have GWAS data available. Stroke subtyping was performed according to the Trial of Org 10172 in Acute Stroke Treatment (TOAST) classification. Detailed phenotypic patient data were recorded in the study database. All subjects are of white, European origin. All the cases have provided a blood sample for the genetic analyses and have been genotyped on the Illumina HumanCoreExome or Illumina Global Screening array. Altogether 558 cases aged 18 to 59 years were available for the present analysis. The study has been approved by the Ethics Committee of Medicine, Helsinki University Hospital. All the participants or their legal representatives have signed an informed consent. Control samples were obtained from the national FINRISK study 1992, 1997, 2002, 2007 and 2012 cohorts .^7^ All the stroke free individuals aged 18 to 59 years at examination, residing in the same geographic area (Greater Helsinki region) and who were followed up until at least 60 years of age were considered as eligible controls.

**INTERSTROKE**

INTERSTROKE (African Americans) is an international, multi-centered, case-control study of stroke investigating the global burden of risk factors across 32 countries and 18 different ethnic groups around the world. Briefly, cases were patients with acute first stroke (within 5 days of symptoms onset and 72 hours of hospital admission) in whom neuroimaging (CT or MRI) was performed. The TOAST classification system was used to define ischemic stroke subtypes. Cases were excluded if 1) they were unable to communicate due to severe stroke without a valid surrogate respondent (e.g. first-degree relative or spouse), 2) they were hospitalized for acute coronary syndrome/myocardial infarction, or 3) stroke was attributed to non-vascular causes (e.g. tumor).

Controls were selected from the community and had no history of stroke. The study was approved by the ethics committees in all participating centres. All participants, or their proxy, provided written informed consent before taking part in the study.

**Ischemic Stroke Genetics Study (ISGS)**

The ISGS is a multicenter prospective, hospital-based inception cohort study of first-ever ischemic stroke. Enrollment for ISGS began in December 2002 and was completed in July 2007. All cases required meeting the WHO definition for stroke and head imaging, by either head MRI or CT, confirmed no alternative cause for the stroke symptoms other than focal cerebral ischemia. All participants had to be over age 18 years. There were no eligibility criteria based on stroke severity or enrollment status in a treatment trial. Cases were excluded if they had CADASIL, MELAS, homocystinuria, or sickle cell anemia or if their stroke was due to vasculitis, vasospasm due to subarachnoid hemorrhage, mechanical aortic valve or mechanical mitral valve, or occurred within 30 days of a vascular surgical procedure. Baseline assessment of patients included standardized assessment of demographics; medical history; vital signs; results of baseline blood tests, pre-stroke functional status per modified Rankin Scale; and National Institutes of Health Stroke Score (NIHSS) by certified examiner. Functional outcomes at 90 days post stroke onset were assessed using telephonic structured interview to obtain Oxford Handicap Scale, Glasgow Outcome Scale and Barthel Index. To minimize center-to-center variability, a single vascular neurologist (Robert D. Brown, Jr., MD) reviewed all available records of every ischemic stroke case for purposes of classification by etiology and syndrome using the TOAST criteria, along with the Baltimore-Washington criteria and the Oxford Community Stroke Project criteria. Medical records were received from the five centers and were stripped of personal identifiers, coded with study ID, and compiled in standard fashion. A separate neurologist independently reclassified all cases using the CCS system.

656 cases of first-ever ischemic stroke and 648 stroke-free controls were enrolled across the 5 centers (Mayo Clinic, Jacksonville, FL, U.S.A.; Mayo Clinic, Rochester, MN, U.S.A.; University of Virginia, Charlottesville, VA, U.S.A.; Shands Hospital, Jacksonville, FL, U.S.A.; Grady Hospital, Atlanta, GA, U.S.A.).

**Krakow**

All consecutive subjects with ischemic stroke (fulfilling WHO criteria) who were admitted to the Stroke Unit at the Jagiellonian University (Krakow, Poland) and who provided informed consent were included in the study. The Stroke Unit serves as a stroke emergency center for one district of Krakow, Poland (200,000 inhabitants) and as a referral center for South East Poland (up to 15% of all admissions). For this on-going, prospective single-center, hospital-based study participants with first ever or recurrent strokes were recruited from January 22, 2002 to September 9, 2010. The Jagiellonian University Bioethics Committee approved the study. Participants in treatment trials were excluded. All subjects were of European origin. Stroke severity was not a criterion for inclusion or exclusion. All cases had performed clinically relevant diagnostic workup, including brain imaging with CT (100%) and/or MRI (up to 20%) as well as ancillary diagnostic investigations including duplex ultrasonography of the carotid and vertebral arteries (approximately 90%), and transthoracic echocardiography (approximately 70%). Magnetic resonance angiography (MRA), computed tomographic angiography (CTA), and ambulatory ECG monitoring, transesophageal echocardiography and blood tests for hypercoagulability were performed. Stroke cases were classified into etiologic subtypes according to TOAST. All cases were phenotyped independently by two experienced stroke neurologists with review of original imaging. Cases were subsequently classified additionally using the CCS system.

The control group included unrelated subjects taken from the population of southern Poland. Control subjects had no apparent neurological disease based on the findings in a structured questionnaire and a neurological examination. The Jagiellonian University Bioethics Committee approved the study and informed consent was obtained from all participants.

**Leuven Stroke Genetics Study (LSGS)**

Cases of European descent with cerebral ischemia, defined as a clinical stroke with imaging confirmation or a TIA with a new ischemic lesion on diffusion-weighted imaging, who were admitted to the Stroke Unit of the University Hospitals (Leuven, Belgium) were enrolled in the LSGS between 2005 and 2009. All participants from the LSGS study underwent brain imaging (MRI in 91% of patients, CT in the remainder) and a standardized protocol including lab examination, carotid ultrasound or CTA and cardiac examination (echocardiography and ambulatory ECG monitoring) in all patients. Based on clinical presentation and results from the diagnostic work-up, cases were classified into ischemic stroke etiologic subtypes according to modified TOAST criteria10 by a single reviewer. Large-vessel disease was defined as either occlusive or significant stenosis (corresponding to > 50% luminal diameter reduction according to North American Symptomatic Carotid Endarterectomy Trial (NASCET) criteria of a clinically relevant pre-cerebral or cerebral artery, presumably due to atherosclerosis. In case CTA was used as the primary imaging modality, stenosis was confirmed by carotid ultrasound. In case of posterior circulation infarcts on imaging, CTA or MRA was used as the primary imaging modality to determine the degree of stenosis. Probable causes of cardiac embolism were excluded.

Cardioembolic stroke was defined as ischemic stroke in the presence of atrial fibrillation, sick sinus syndrome, myocardial infarction in the past four weeks, cardiac thrombus, infective endocarditis, atrial myxoma, prosthetic mitral or aortic valve, valvular vegetations, left ventricular akinetic segment, dilated cardiomyopathy, or patent foramen ovale or atrial septal aneurysm. Significant stenosis/occlusion due to atherosclerosis of an appropriate pre-cerebral or cerebral artery should be excluded. Other determined cause of stroke included those with arterial dissection, vasculitis, hematologic disorders, monogenic syndromes, and complications of cardiovascular procedures. Dissection was diagnosed by typical findings on contrast-enhanced MRA and T1-fat suppressed MRI. Cryptogenic stroke was defined when no cause was identified despite an extensive evaluation. Strokes associated with significant aortic arch atheroma with plaques of ≥ 4 mm were also considered cryptogenic strokes. In addition to this primary classification, cases were reclassified using CCS.

Control individuals were recruited in the same population amongst healthy individuals, spouses of patients suffering from neurological diseases (amyotrophic lateral sclerosis, ischemic stroke, or multiple sclerosis), and from the Leuven University Gerontology Database as previously described.

**Lund Stroke Register (LSR)**

The LSR is an ongoing study including consecutive subjects with first-ever stroke since March 1, 2001 from the local uptake area of Skåne University Hospital, Lund (Sweden). Stroke was defined using the WHO criteria. Subjects aged 18 years or older with stroke caused by cerebral infarct, intracerebral hemorrhage or subarachnoid hemorrhage are included. Cases are included regardless of stroke severity, race-ethnic group belonging, or participation in any treatment trial. Those with iatrogenic or traumatic stroke are excluded. In the discovery phase of the SiGN study, subjects from LSR with first-ever ischemic stroke between March 1, 2001 and February 28, 2010 were included if they or their next of kin provided informed consent. Age over 90 years was set to 90 years to maintain anonymity. Every participant underwent CT, MRI, or autopsy of the brain, and ECG. Echocardiography, ultrasound, CTA or MRA of cerebral arteries was performed when judged clinically relevant. The subtype of ischemic stroke was determined using CCS.

For the secondary phase of SiGN, LSR individuals not included in the SiGN discovery phase participated after genotyping in the South Swedish genome-wide association study as follows: first ever ischemic stroke cases recruited in 2006 and 2010 to 2012, and age- and sex-matched LSR control subjects without stroke recruited in 2001 to 2002 and 2006 to 2007 from the same geographical area with use of the official Swedish population register.

**Middlesex County Ischemic Stroke Study (MCISS)**

The MCISS was initiated as a prospective hospital-based stroke registry at the New Jersey Neuroscience Institute (Edison, NJ, U.S.A.). All cases over age 18 years were included, and no specific ethnic/racial group was targeted or excluded. From 2000 to 2009, 1,139 subjects with ischemic strokes were enrolled in this registry. There was no selection criterion based upon stroke severity, and both first-ever and recurrent strokes were included. Cases that were participants in treatment trials were not excluded. The major race/ethnic groups are Whites (67.2%), African Americans (14.3%), Asian Indians (8.2%), Hispanic (5.5%) and others (4.8%, Chinese and other Asians). All subjects with clinical suspicion of a stroke were admitted through the emergency room to a dedicated stroke unit supervised by a vascular neurologist. After a history and neurological examination, a standardized series of investigations were performed: complete blood count and differential, comprehensive metabolic panel, electrolytes, blood urea nitrogen, creatinine, lipid panel (total cholesterol, low-density lipoprotein, high-density lipoprotein, triglyceride levels, homocysteine levels, a cerebral MRI/MRA (if the MRI could not be performed, a head CT scan was done), carotid duplex ultrasound, ECG and an echocardiogram. The diagnosis of cerebral infarct was confirmed by the imaging studies. The epidemiological and clinical data on these participants was collected prospectively. Two independent investigators (one of which was a board-certified neurologist with expertise in vascular neurology) reviewed the data, and all strokes were classified into etiological subtypes using TOAST criteria. In addition, the Oxfordshire stroke classification was applied, and the vascular distribution of stroke was tabulated. All procedures, including the generation of the databases and recruitment of the stroke subjects, were conducted following Institutional Review Board policies and procedures at the New Jersey Neuroscience Institute/JFK Hospital.

Inclusion criteria was all subjects over the age of 18 years with clinical suspicion of a stroke. Exclusion criteria was subjects under the age of 18 years.

A control cohort of stroke free patients was also established. These volunteers have been recruited from the offices of local primary care physicians in Middlesex County, New Jersey and from the neurology clinic based at the New Jersey Neuroscience Institute. In addition to the clinical information, DNA samples have been obtained from blood samples for the control groups.

**Miami Stroke Registry and Biorepository (MIAMISR)**

The MIAMISR at the University of Miami/Jackson Memorial Hospital (Miami, FL, U.S.A.) is an ongoing prospective hospital registry of consecutive patients subjects with prevalent stroke (ischemic and hemorrhagic) and TIA with available neuroimaging (CT or MRI) who provide informed consent. There are no specific exclusion criteria with the respect to age, stroke severity, disability or participation in treatment trials. It was established in November of 2008 in order to investigate stroke type, ischemic stroke subtypes, stroke genetics and stroke outcomes in diverse ethnic population of Miami. The stroke population is predominately Hispanic (63%), with Cuba (32%), Nicaragua (4.8%), Colombia (4.8%), and Puerto Rico (4.1%) contributing the most subjects. Jackson Memorial Hospital is a 1,550-bed county hospital affiliated with the University of Miami with approximately 900 stroke and TIA admissions per year. Demographic and clinical data along with blood samples for genetic and other research have been collected prospectively during the hospitalizations. Follow-up information was obtained at 90 days by telephone interview or in person. Trained research staff obtained written informed consent from the stroke patients or the health care proxy when available for participation in MIAMISR.

**Milano**

This study includes consecutive Italian patients referred to Besta Institute from 2000 to 2009 with stroke and included in the Besta Cerebrovascular Diseases Registry (CEDIR). Ischemic stroke cases, first ever or recurrent, confirmed on brain imaging, were selected for this study. All cases were of self-reported Caucasian ancestry and had clinically relevant diagnostic workup performed. All cases were phenotyped by an experienced stroke neurologist according to TOAST criteria, based on relevant clinical imaging and available information on cardiovascular risk factors. Controls are Italian individuals enrolled within the PROCARDIS Study, with no personal or sibling history of coronary heart disease before age 66 years.

**Munich**

Subjects with first-ever or recurrent ischemic stroke were recruited consecutively from a single dedicated stroke unit (University Hospital, LMU Munich, Munich, Germany) from 2002 onward. All participants were over the age of 18 years and of European descent. Brain imaging was performed in all cases, with most patients (> 80%) undergoing MRI, including DWI. Diagnosis of ischemic stroke was based on neurological symptoms in combination with a documented acute infarct on neuroimaging. Subjects were not excluded based on stroke severity or whether they were enrolled in a treatment trial. Diagnostic workup included ECG and duplex ultrasonography of the extracranial carotid arteries in all cases. Transcranial ultrasonography, CTA and/or MRA, transthoracic and transesophageal echocardiography, and ambulatory ECG were performed if clinically indicated.

For the German MUNICH discovery samples and the Stroke in Young Fabry Patients (SIFAP) samples, independent control groups were selected from Caucasians of German origin participating into the population KORAgen study. This survey represents a sex- and age stratified random sample of all German residents of the Augsburg area and consists of individuals 25 – 74 years of age, with about 300 subjects for each 10-year increment. All controls were free of a history of stroke or transient ischemic attack.

**Nurses’ Health Study**

The NHS cohort consists of 121,700 female registered nurses aged 30 – 55 years who were residing in 11 U.S. states and who were enrolled in 1976 through responding to a mailed questionnaire on their medical history and lifestyle practices. They have been followed with biennial mailed questionnaires collecting information on disease risk factors and health status. From 1989 – 1990, blood samples were collected from 32,826 participants. Among these participants, we prospectively identified incident strokes and confirmed ischemic stroke cases by medical record review. Clinical symptoms consistent with stroke and exclusion of alternate etiologies were required for classification of stroke. Virtually all cases had imaging, but confirmation on CT or MRI was not required. No participants were excluded based on race/ethnicity. Neither stroke severity nor enrollment in a treatment trial was part of the eligibility criteria.

Incident ischemic strokes that were part of R01 biomarkers for ischemic stroke were included if sufficient clinical data was available.

**Northern Manhattan Study (NOMAS)**

NOMAS is an ongoing population-based study designed to determine stroke incidence, risk factors and outcome in an urban multiethnic population. NOMAS started in 1993 as a case-control study of index ischemic stroke cases admitted to the Columbia University Presbyterian Medical Center (New York, NY, U.S.A.) and affiliated hospitals and matching community controls (Northern Manhattan Stroke Study, NOMASS) and continued as a prospective stroke incidence study by following up controls in 1997 (NOMAS). Demographic and clinical data were collected prospectively during the hospitalizations and annually by phone or in person. Genetic samples were derived from two sources: (a) the population-based case-control study conducted from 1993-98 (NOMASS) and (b) the ongoing prospective cohort study (NOMAS). First-ever ischemic stroke cases were identified for the case control study by screening of patient admissions, discharge codes, and referrals for neuroimaging at 15 acute care hospitals in the defined study area and multiple approaches to monitor for non-hospitalized cases. Incident ischemic stroke cases were identified from the prospective cohort study through follow-up visits and scheduled telephone contacts. Ischemic stroke cases from both sources were followed at 6 months by telephone and then annually afterwards in order to assess functional status and other outcomes. The administrative coordinating center of NOMAS moved from New York to Miami in 2007. The Institutional Review Boards of both institutions, Columbia University, and the University of Miami (Miami, FL, U.S.A.), approved the study.

NOMAS started in 1993 as a case-control study of index ischemic stroke cases admitted to the Columbia University Presbyterian Medical Center (New York, NY, U.S.A.) and affiliated hospitals and matching community controls (Northern Manhattan Stroke Study, NOMASS) and continued as a prospective stroke incidence study by following up controls in 1997 (NOMAS).

**Odyssey**

ODYSSEY is a multicenter prospective cohort study on the prognosis and risk factors of patients with a first-ever TIA, ischemic stroke or intracerebral hemorrhage aged 18 to 50 years among 1490 patients. Primary outcome includes all-cause mortality and risk of recurrent vascular events. Secondary outcome will be the risk of post-stroke epilepsy and cognitive impairment. Patients have completed structured questionnaires on outcome measures and risk factors. Both well-documented and less well-documented risk factors and potentially acute trigger factors will be investigated. Patients will be followed for at least 10 years.

**OXVASC**

OXVASC is an on-going population-based study of the incidence and outcome of cerebrovascular, cardiovascular, and peripheral vascular events since April 1, 2002. The OXVASC study population comprises all 91,105 individuals, irrespective of age, registered with 101 general practitioners in 9 general practices in Oxfordshire, UK. Multiple overlapping methods of “hot” and “cold” pursuit are used to achieve near complete ascertainment of as many cases as possible. All subjects are consented and seen by study physicians as soon as possible after their initial presentation. In the SiGN study, cases of all ethnic groups from OXVASC with any ischemic stroke between April 1, 2002 and August 31, 2010 were included if they consented to have research DNA samples extracted. Ischemic stroke was defined as an episode of focal neurological deficits with acute onset lasting > 24 hours or until death, with no apparent non-vascular cause, and no signs of primary hemorrhage on brain imaging. An infarct did not need to be seen on CT or MRI to be included in this study. Cases were not excluded if they were of a treatment trial or for their stroke severity. Demographic data, major vascular risk factors (hypertension, diabetes, smoking, hyperlipidemia, prior TIA and history of coronary disease or peripheral vascular disease), and symptomatology were recorded in all patients. Cases routinely had brain imaging (CT or MRI), vascular imaging (carotid Doppler or CTA /MRA or digital subtraction angiography), and 12-lead ECG. Echocardiography and 24-hour ambulatory ECG monitoring were done in selected patients. A senior neurologist subsequently reviewed all cases, and stroke etiology was classified according to modified TOAST criteria. Risk factors such as hypertension and diabetes were not included in the criteria. The subjects were classified as undetermined stroke only if the diagnostic workup was complete (any form of brain imaging plus ECG and any form of vascular imaging), but no clear etiology was found. Those with incomplete investigation were classified as unknown stroke while stroke of multiple causes was classified separately. Controls are collected through spouse or friend volunteers identified by TIA/stroke patients.

**The Risk Assessment of Cerebrovascular Events Study (RACE 1 & RACE 2)**

The Risk Assessment of Cerebrovascular Events (RACE) Study is a retrospective case-control study designed to identify and evaluate genetic, lifestyle and biomarker determinants of stroke and its subtype in Pakistan. Samples were recruited from six hospital centres in Pakistan. Cases were eligible for inclusion in the study if they: (1) are aged at least 18 years; (2) presented with a sudden onset of neurological deficit affecting a vascular territory with sustained deficit at 24 hours verified by medical attention within 72 hours after onset (onset is defined by when the patient was last seen normal and not when found with deficit); the diagnosis was supported by CT/MRI; and (4) presented with a Modified Rankin Score of < 2 prior to the stroke. TOAST and Oxfordshire classification systems were used to sub-phenotype all stroke cases.

Control participants were individuals enrolled in the Pakistan Risk of Myocardial Infarction Study (PROMIS), a case/control study of acute MI based in Pakistan.^8^ Controls in PROMIS were recruited following procedures and inclusion criteria as adopted for RACE cases. In order to minimize any potential selection biases, PROMIS controls selected for this stroke study were frequency matched to RACE cases based on age and gender and were recruited in the following order of priority: (1) non-blood related or blood related visitors of patients of the out-patient department; (2) non-blood related visitors of stroke patients; (3) patients of the out-patient department presenting with minor complaints.

**Reasons for Geographic and Racial Differences in Stroke (REGARDS)**

The REGARDS study is a U.S. national, population-based, longitudinal cohort of 30,239 African American and white adults aged ≥ 45 years, recruited January 2003 to October 2007 with ongoing follow-up. Suspected stroke is queried every six months and triggered by participant self-report of stroke, stroke symptom(s), hospitalization, or proxy report of death. Stroke severity and participation in a treatment trial did not limit inclusion in this study. Medical records for these reported events were retrieved and reviewed by at least two members of a committee of stroke experts with disagreements resolved by a third adjudicator. A symptom-based approach, independent of neuroimaging outcome, was used to confirm events using the WHO definition of stroke. An infarct did not need to be seen on brain imaging to be included in this study. Ischemic stroke subtype classification was conducted using the TOAST system.

**Sahlgrenska Academy Study on Ischemic Stroke (SAHLSIS)**

SAHLSIS is a case-control study of ischemic stroke based in Gothenburg, Sweden. Adult subjects who presented with first-ever or recurrent acute ischemic stroke before 70 years of age were recruited consecutively at stroke units in western Sweden from 1998 to 2012. All participants were of European origin. Patients were not excluded based on stroke severity or whether they were enrolled in a treatment trial. All participants underwent ECG and neuroimaging at the acute stage (all by CT and 58% also by MRI). Additional diagnostic work-up was performed as clinically indicated. Inclusion criteria was ischemic stroke which was defined as an episode of focal neurological deficits with acute onset and lasting > 24 hours or until death, with no apparent non-vascular cause, and no signs of primary hemorrhage on brain imaging. Subjects were excluded if they had a diagnosis of cancer at advanced stage, infectious hepatitis, or human immunodeficiency virus. Ischemic stroke was assigned according to modified TOAST criteria.

Healthy Caucasian community controls (n=600) matched for age and sex from the same geographic area were randomly selected from participants in a population-based survey or the Swedish Population Register.

**Stroke in Young Fabry Patients (SIFAP)**

The SIFAP study is a multicenter study carried out to determine the frequency of Fabry disease in an unselected group of young adult patients with acute cerebrovascular events defined as having had an acute ischemic stroke or transient ischemic attack less than three months before enrollment into the study. First-ever (80.5%) and recurrent ischemic strokes were included. MRI was a mandatory procedure but, in the case of negative or missing MRI, a qualified stroke neurologist could confirm the clinical diagnosis. For this project, ischemic stroke cases recruited from 15 sites throughout Germany and determined not to have Fabry Disease were included in the analysis. All were of European ancestry and had age of first stroke of 18 – 55 years. The diagnosis of Fabry disease was based in males as well as in females in the first level on the sequencing data of the entire exon structure including promoter of the α-galactosidase gene. In cases where a mutation was detected, biochemical analysis was done. Stroke cases from SIFAP were genotyped at CIDR (Baltimore, MD) using the Illumina Human Omni 2.5MQuad array. Only those cases without Fabry disease were selected for genotyping.

Controls free of cardiovascular diseases were selected from the KORA Study previously genotyped at CIDR in the same platform. The Cooperative Health Research in the Region of Augsburg (KORA) study is a population-based study of cardiovascular and metabolic traits carried out in the region of Augsburg, Southern Germany. A subset of control subjects (N = 28) was re-genotyped together with cases to provide cross-set duplicates. This joint clustering was used to minimize possible artifactual differences in allelic frequency between cases and controls due to genotyping at different times, and the cross-set duplicates were used to detect such artifacts that may have occurred.

**The South London Ethnicity and Stroke Study (SLESS)**

The South London Ethnicity and Stroke Study (SLESS) is a prospective study begun in 1999 that has recruited consecutive black patients with stroke from a contiguous catchment area covered by 3 hospitals in South London (Guy’s and St Thomas’ Hospitals, King’s College Hospital, and St George’s Hospital). Ethnicity was defined according to the UK Census 2001 definition and classified as Black African or Black Caribbean. The study was reviewed and approved by the Wandsworth Local Research Ethics Committee, and informed consent was obtained from all participants. One consultant neurologist performed stroke subtyping using data collected on a standard proforma with additional review of all original brain imaging in all patients, as well as review of original notes when necessary. The pathophysiological Trial of Org 10172 in Acute Stroke Treatment (TOAST) subtyping classification was used for subtyping of ischemic stroke. Stroke ascertainment in SLESS: One consultant neurologist performed stroke subtyping using data collected on a standard proforma with additional review of all original brain imaging in all patients, as well as review of original notes when necessary. The pathophysiological Trial of Org 10172 in Acute Stroke Treatment (TOAST) subtyping classification was used for subtyping of ischemic stroke.

Recruitment of black controls was done by random selection from General Practice lists in the catchment areas of St George’s, Guys and St Thomas, and King’s College Hospital between 1999 and 2012. Potential controls were selected from age and gender strata matched to stroke cases. Furthermore, controls were identified within St George’s University of London and St George’s Hospital staff and contacted via email. Additionally, posters inviting healthy Black African and Black Caribbean individuals were displayed in local leisure centres, General Practice surgeries, churches, and community centres within the same catchment area as the that of the cases.

**Secondary Prevention of Small Subcortical Strokes (SPS3)**

The SPS3 trial (NCT00059306) is a randomized, multicenter, Phase 3 trial of antiplatelet therapy and antihypertensive therapy. Participants are randomized to aspirin alone or the combination of aspirin and clopidogrel. Participants are also randomized to two groups of blood pressure control: either to a target systolic blood pressure of 130 – 149 mm Hg or < 130 mm Hg. Principal eligibility criteria include men or women at least 30 years of age with clinical evidence of small subcortical stroke and brain MRI evidence of small subcortical infarct. Subjects were required to not have evidence of ipsilateral symptomatic cervical carotid stenosis or high-risk cardioembolic sources for embolism. Primary outcomes included ischemic and hemorrhagic stroke. DNA samples were collected from 38% (1,139/3,020) of participants in the trial. These samples were obtained from 46% (37/81) participating centers across the U.S., Canada, Spain, Mexico, Chile, Ecuador, and Peru. No additional eligibility criteria were necessary beyond informed consent for participating in the DNA sub-study.

**St. George**

First-ever and recurrent ischemic stroke cases of European descent attending a cerebrovascular service were recruited from 1995 to 2008. All cases were phenotyped by one experienced stroke neurologist with review of original imaging. All participants had clinically relevant diagnostic workup performed, including brain imaging with CT and/or MRI as well as ancillary diagnostic investigations including duplex ultrasonography of the carotid and vertebral arteries or MRA/CTA, blood tests, and ECG, and where clinically indicated echocardiography and ambulatory ECG monitoring was performed. Cases were enrolled only if a symptomatic acute infarct was detected on head imaging. Participants had to be over the age of 18 years and have provided informed consent. No case was excluded for participation in a treatment trial or because of stroke severity. An algorithm was established to use the clinical trials database to automatically populate the web-based CCS tool to generate CCS stroke subtype diagnoses. St. George cases were combined with DNA Lacunar Plus (removing any duplicates) and utilized the same set of controls.

**Siblings with Ischemic Stroke Study (SWISS)**

SWISS is a prospective, hospital-based affected sibling pair study of ischemic stroke. Enrollment for SWISS began in December 2000 and was completed in February 2011. DNA samples were collected from 312 ischemic stroke-affected sibling pairs. During this time, 1,026 cases with first-ever or recurrent ischemic stroke were enrolled across 70 centers in North America (66 in the U.S. and 4 in Canada). All probands required at least one living sibling with a history of stroke and required meeting the WHO definition for stroke with head imaging, by either head MRI or CT, confirming no alternative cause for the stroke symptoms other than focal cerebral ischemia. Probands were excluded if they had CADASIL, MELAS, homocystinuria, or sickle cell anemia or if their stroke was due to vasculitis, vasospasm due to subarachnoid hemorrhage, mechanical aortic valve or mechanical mitral valve, or occurred within 30 days of a vascular surgical procedure. Baseline assessment of cases included standardized assessment of demographic and medical history. Siblings were recruited primarily using proband-initiated contact. Stroke-affected siblings were screened using the Questionnaire for Verifying Stroke-free Status (QVSS). Eligibility criteria for affected siblings were the same as for probands. The Stroke Verification Committee, composed of two vascular neurologists, confirmed ischemic stroke status in affected siblings by medical record review. For probands, the center principal investigator classified ischemic stroke using the original TOAST classification system. Center principal investigators were neurologists certified in TOAST classification using stroke vignette training and certification process. The Stroke Verification Committee classified all affected siblings using TOAST based on medical record review. The Committee received medical records stripped of personal identifiers, coded with study identification number, and compiled in standard fashion. All participants gave written informed consent for participation in the study, and the local institutional review boards of each individual clinical center and the Mayo Clinic institutional review board approved the study. Discordant siblings of the proband were confirmed to be stroke-free using the Questionnaire for Verifying Stroke-free Status.

**UK Biobank Stroke Study**

The UK Biobank (UKB) <https://www.ukbiobank.ac.uk/> was established to improve understanding of common diseases including stroke. Participants were recruited from the general adult population and, in addition to having provided self-reported medical history at recruitment, are followed prospectively, chiefly through linkage to their National Health Service records.

For definition of stroke cases, all ischemic stroke cases from the algorithmically defined stroke outcomes (UKB fields 42008 and 42990) were used. If there was no date of stroke defined, this case was removed from the analysis. Furthermore, cases with an incident hemorrhagic stroke before the index ischemic stroke were also removed from the analysis. Only individuals of White British ancestry, as defined by UK Biobank, were selected.

Controls were all individuals in the UKB of White British Ancestry that had no history of any type of stroke (including both ischemic and hemorrhagic stroke).

**VHIR-FMT- Barcelona**

The Barcelona cohort is a subset of Caucasian ischemic stroke subjects that were enrolled as a part of the Genetic contribution to functional Outcome and Disability after Stroke (GODs) project. Cases were selected through demonstration of acute ischemic stroke in a neuroimaging study during the first 7 days after stroke. We included cases with a first-ever and with a recurrent stroke. We did not include lacunar strokes due to the study was focused only on disability after stroke of non-lacunar cases. We did not use age or stroke severity as exclusion criteria. Participants were not part of a treatment trial. Etiologic subgroups were classified following TOAST criteria. All the samples (cases and controls) were genotyped using the Infinium Human Core Exome Chip (Illumina). Written informed consent was obtained from all subjects with approval from the ethics committee of all participating institutions.

The control cohort was collected in primary care centers from Barcelona city and some hospital in the Spanish Network as a part of the Investigating Silent Stroke in hYpertensives: A magnetic resonance imaging Study (ISSYS) and Genotyping RECurrence Risk of Stroke (GRECOS) study. Controls were healthy subjects older than 40 years old without history of ischemic stroke.

**Vitamin Intervention for Stroke Prevention (VISP)**

The VISP trial was a multicenter, randomized double-blind controlled clinical trial that enrolled subjects aged 35 years or older with homocysteine levels above the 25th percentile at screening and a non-disabling cerebral infarction within 120 days of randomization. Non-disabling cerebral infarction was defined as an ischemic brain infarction not due to embolism from a cardiac source, characterized by the sudden onset of a neurological deficit. The deficit must have persisted for at least 24 hours, or, if not, an infarction in the part of the brain corresponding to the symptoms must have been demonstrated by CT or MRI. The trial was designed to determine if daily intake of a multivitamin tablet with high-dose folic acid, vitamin B6 and vitamin B12 reduced recurrent cerebral infarction (primary endpoint), and nonfatal myocardial infarction or mortality (secondary endpoints). Subjects were randomly assigned to receive daily doses of the high-dose formulation (N = 1,827), containing 25mg pyridoxine (B6), 0.4mg cobalamin (B12), and 2.5mg folic acid; or the low-dose formulation (N = 1,853), containing 200µg pyridoxine, 6µg cobalamin and 20µg folic acid. Enrollment began in August 1997 and ended in December 2001, with 3,680 participants enrolled, from 55 clinic sites across the US and Canada and one site in Scotland. All participants provided written informed consent, and all local governing institutional review boards approved the trial. A subset of VISP participants provided separate consent for genetic analyses.

Control data for comparison with European ancestry VISP stroke cases were obtained through the database of genotypes and phenotypes (dbGAP) High Density SNP Association Analysis of Melanoma: Case-Control and Outcomes Investigation (phs000187.v1.p1; R01CA100264, 3P50CA093459, 5P50CA097007, 5R01ES011740, 5R01CA133996, HHSN268200782096C; PIs Christopher Amos, Qingyi Wei, Jeffrey E. Lee). For VISP stroke cases of African ancestry, a subset of the Healthy Aging in Neighborhoods of Diversity across the Life Span study (HANDLS) were used as stroke free controls.

**Women’s Health Initiative Observational Study (WHI-OS)**

The Women’s Health Initiative Observational Study (WHI-OS) is a long-term follow-up study of postmenopausal women to identify and assess the effects of biological, genetic and lifestyle risk factors for cancer, cardiovascular disease, osteoporosis and other diseases of older women. The cases submitted here came from a case-control ancillary study nested within the WHI-OS of the first 972 strokes occurring after WHI-OS baseline. This case-control study was the Hormones and Biomarkers Predicting Stroke Study (HaBPS), conducted to examine blood biomarkers in relation to stroke. Forty clinical centers throughout the United States enrolled 93,676 women ages 50 to 79 years at baseline into the parent study, the WHI-OS, between September 1993 and February 28, 1997. Follow-up for clinical events and exposures is ongoing. Recruitment into WHI-OS was mostly through mass mailings to age-eligible women from large mailing lists such as voter registration, driver’s license, Health Care Financing Administration, or other insurance lists. Recruitment of minorities and older women was a particular study objective. Women were either specifically recruited for the Observational Study or entered it because they were ineligible or unwilling to be randomized into the Women’s Health Initiative Clinical Trials of hormone therapy and/or dietary modification. Exclusions from WHI-OS were participation in other randomized trials, predicted survival of < 3 years, alcoholism, drug dependency, mental illness, dementia, or other conditions making them unable to participate in the study. Exclusions for the HaBPS case-control study of biomarkers of stroke were women with prior history of myocardial infarction or stroke or those who did not have adequate blood sample for biomarker assays. Strokes were first identified through annual mail and/or telephone follow-up, and participant or third-party reports of overnight hospitalizations which were further investigated by obtaining laboratory results, medical records, and available imaging study reports. Trained local physician adjudicators assigned a diagnosis according to standard criteria. Locally adjudicated strokes were sent for central adjudication by three neurologists. Two neurologists adjudicated each potential case, and disagreements were resolved by conference call consensus of the three neurologists. Only centrally confirmed ischemic strokes that required hospitalization were used in this study. TIAs and hemorrhagic strokes (determined on review of reports of brain imaging studies) were excluded. Ischemic stroke was defined as the rapid onset of a persistent neurologic deficit attributed to a vessel occlusion lasting more than 24 hours and without evidence for other causes. The deficit must have lasted > 24 hours unless death supervened or there was a lesion compatible with acute stroke demonstrated on CT or MRI scan. Ischemic strokes were also centrally classified by TOAST and CCS criteria.

**Washington University St. Louis (WUSTL) Study**

The WUSTL patient collection included ischemic stroke cases admitted to Barnes-Jewish Hospital/Washington University Medical Center (St. Louis, MO, U.S.A.) for genetic studies starting from August 1, 2008. Participants were identified for the genetic studies by screening admissions at our tertiary care hospital (both in the Emergency Department and on the Inpatient Stroke Service) without regard to age, race or ethnicity, including both first-ever and recurrent strokes. Subjects were retained in the study if their discharge diagnosis was ischemic stroke (without requirement for the stroke to be visualized on CT or MRI). Demographic and clinical data were collected prospectively during the hospitalization and at 90 days, by phone or in person. Genetic samples were derived from subjects enrolled in 3 different studies: (a) Acute tPA pharmacogenomics study (Ischemic stroke cases who received tPA and were admitted to BJH/Washington University; serial NIHSS scores, and data on hemorrhagic transformation was collected) (b) Recovery Genomics after Ischemic Stroke Study (ReGenesIS, Ischemic stroke cases with NIHSS > 3 points without underlying chronic neurological disease, and expected survival up to 3 months after stroke), and (c) the Cognitive Recovery and Rehabilitation Group (CRRG) Registry (all ischemic stroke cases admitted to BJH/Washington University who consent to entering their clinical data into a stroke registry, and the collection of blood for genetic analysis). Cases that were part of a treatment trial were excluded from the tissue plasminogen activator pharmacogenomics and ReGenesIS study, but not the CRRG registry.

Stroke subtyping using TOAST criteria/etiologies was done for all subjects participating in the studies. A subset of subjects was further subtyped according to the CCS (Causative Classification System of Ischemic Stroke) as part of the SiGN study.

**Young Lacunar Stroke DNA Resource Plus**

A total of 1,029 Caucasian patients with lacunar stroke, aged ≤70 years, were recruited from 72 specialist’s stroke centres throughout the UK between 2002 and 2012, as part of the Young Lacunar Stroke DNA Resource. DNA samples were available in 930 patients. An additional 82 Caucasian patients of all ages with lacunar stroke were recruited from St. George’s Hospital, London as part of the GENESIS study. Lacunar stroke was defined as a clinical lacunar syndrome, with an anatomically compatible lesion on MRI (subcortical infarct ≤15 mm in diameter). All patients underwent full stroke investigation including brain MRI, imaging of the carotid arteries and ECG. Echocardiography was performed when appropriate. All MRIs and clinical histories were reviewed centrally by one physician. Exclusion criteria were: stenosis >50% in the extra- or intracranial cerebral vessels, or previous carotid endarterectomy; cardioembolic source of stroke, defined according to the TOAST (Trial of Org 10172 in Acute Stroke Treatment) criteria as high or moderate probability; cortical infarct on MRI; subcortical infarct > 15mm in diameter, as these can be caused by embolic mechanisms (striatocapsular infarcts); any other specific cause of stroke (e.g. lupus anticoagulant, cerebral vasculitis, dissection, monogenic cause of stroke). All cases were screened for *NOTCH3* CADASIL and Fabry disease mutations and positive cases excluded. All patients and controls underwent a standardized clinical assessment and completed a standardized study questionnaire. MRI was not performed in controls. The study was approved by the Multi-Centre Research Ethics Committee (04/MRE00/36) and informed consent was obtained from all participants.

Unrelated Caucasian controls, free of clinical cerebrovascular disease, were obtained by random sampling, stratified for age and sex, from general practice lists from the same geographical location as the patients.

**Control-Only Cohorts:**

**Attention-deficit Hyperactivity Disorder (ADHD)**

The Vall d'Hebron Research Institute (VHIR) cohort included 435 blood donors of Caucasian origin recruited from 2004 to 2008 at the Hospital Universitari Vall d'Hebron (Barcelona, Spain) to identify loci conferring susceptibility to Attention-Deficit Hyperactivity Disorder. Seventy-six percent of participants were male (N = 330) and the average age at assessment was 43.8 years (s.d. = 14.3). Genome-wide genotyping was performed with the Illumina HumanOmni1-Quad BeadChip platform. The study was approved by the ethics committee of the institution and informed consent was obtained from all participants in accordance with the Declaration of Helsinki.

**FINRISK**

FINRISK surveys are cross-sectional, population-based studies conducted every 5 years since 1972 to monitor the risk of chronic diseases. For each survey, a representative random sample was selected from 25- to 74-year-old inhabitants of different regions in Finland. The survey included a questionnaire and a clinical examination, at which a blood sample was drawn, with linkage to national registers of cardiovascular and other health outcomes. The current study included eligible individuals from FINRISK surveys conducted in 1992, 1997, 2002, and 2007. The GWAS genotyping has been done earlier in several phases for different substudies: PredictCVD, Corogene and CoreExome. Participants with a history of stroke were excluded. Stroke ascertainment in FINRISK: During follow-up, participants were monitored for stroke through linkage of the study database with the National Hospital Discharge Register and the National Causes-of-Death Register. The clinical outcomes were linked to study subjects using their unique national social security ID, which is assigned to every permanent resident of Finland. The registers are countrywide covering all cardiovascular events that have led either to hospitalization or death in Finland. With both registers the diagnostic classification was done using the Finnish adaptation of ICD-codes: I63; not I63.6, I64 (ICD-10) / 4330A, 4331A, 4339A, 4340A, 4341A, 4349A, 436 (ICD-9) / 433, 434, 436 (ICD-8) for Ischemic stroke excluding any hemorrhagic strokes, and I60-I61,I63-I64 (not I63.6) (ICD-10) / 430, 431, 4330A, 4331A, 4339A, 4340A, 4341A, 4349A, 436 (ICD-9) / 430, 431 (except 431.01, 431.91), 433, 434, 436 (ICD-8) for allstroke including SAH. ICD-8 codes 430, 431 (excluding codes 431.01, 431.91 of the Finnish adaptation of ICD-8*), 432, 433, 434 or with ICD-9 codes 430, 431, 433 (excluding codes 4330X, 4331X, 4339X of the Finnish adaptation of ICD-9*), 434 (excluding code 4349X of the Finnish adaptation of ICD-9*), 436, 437, 438 or with ICD-10 codes I60, I61, I63 (excluding I63.6), I64 or I69.34. An event found in either register was sufficient for diagnosis.

**The Hispanic Community Health Study/Study of Latinos (HCHS/SOL)**

The Hispanic Community Health Study/Study of Latinos (HCHS/SOL) was initiated in 2006 to investigate the prevalence and risk factors affecting several health conditions, including heart, lung and blood disorders, kidney and liver function, diabetes, cognitive function, dental conditions and hearing disorders.^9^ Participants aged 18 – 74 self-identified as Hispanic or Latino, with substantial representation of Mexican, Puerto Rican, Dominican, Cuban, Central and South American groups. They were recruited from four field centers in the United States: San Diego, CA; the Bronx, NY; Chicago, IL; and Miami, FL. 12,803 study participants consented to genetic studies and will be included in the HCHS/SOL dbGaP posting. Genotyping of the HCHS/SOL participants was performed at Illumina Microarray Services using the SOL HCHS Custom 15041502 array (annotation version “B3”, genome build 37), which includes 2,575,443 variants (of which 2,427,090 are in common with the Illumina HumanOmni2.5 and 148,353 are custom content).

**Health and Retirement Study (HRS)**

The University of Michigan Health and Retirement Study (HRS) is a longitudinal panel study that surveys a representative sample of more than 20,000 Americans over the age of 50 every two years. Supported by the National Institute on Aging (NIA U01AG009740) and the Social Security Administration, the HRS explores the changes in labor force participation and the health transitions that individuals undergo toward the end of their work lives and in the years that follow. Since its launch in 1992, the study has collected information about income, work, assets, pension plans, health insurance, disability, physical health and functioning, cognitive functioning, and health care expenditures. HRS is intended to be a nationally representative sample with 2:1 oversampling of minority groups including African American and Hispanic/Latino populations.^10^ In Phases I – II, 12,507 study participants with DNA samples taken in 2006 or 2008 were included in the dbGaP posting. Genotyping of the HRS Phase I – II participants was performed at CIDR using the Illumina HumanOmni2.5-4v1 array (annotation version “D”, genome build 37) and released a total of 2,443,179 variants. Only participants in the European ancestry and African ancestry analytic samples (defined by self-reported race/ethnicity and genetic principal components as described in the dbGaP documentation) were included in this analysis. Participants with self-reported stroke or self-reported heart disease were excluded.

**INfancia y Medio Ambiente (INMA)**

The INfancia y Medio Ambiente (Environment and Childhood) project is a research project comprising a Spanish population-based birth cohort created to study the role of the environmental pollutants during pregnancy and first stages of life and their effects on childhood growth and development. The cohort was established between 2003 and 2008 from mothers enrolled in four regions within Spain and included their infants.

**KORA**

For the German MUNICH discovery samples and the Stroke in Young Fabry Patients (SIFAP) samples, independent control groups were selected from Caucasians of German origin participating into the population-based KORA study. This survey represents a sex- and age stratified random sample of all German residents of the Augsburg area and consists of individuals 25 – 74 years of age, with about 300 subjects for each 10-year increment. All controls were free of a history of stroke or transient ischemic attack. KORA samples were genotyped on the Illumina Human 550k platform. QC was identical for all WTCCC cohorts.

**Malmo Diet and Cancer (MDC) Study**

The MDC study is a population-based prospective cohort study. A total of 30,447 individuals, 45 to 73 years old, 60% women, attended a baseline examination between February 1991 and September 1996. Between 1992 and 1994, a total of 6,103 randomly selected subjects attended an extended baseline examination with the purpose of studying the epidemiology of cardiovascular diseases (the MDCcardiovascular cohort, MDC-CC). At the baseline examination, 23% of the participants were smokers, 16% used anti-hypertensive medication, 14% were obese (body mass index > 30 kg/m^2^), 88% were born in Sweden and > 99% were born in Europe. Genotyping was performed using the Illumina Infinium Omni5 platform with exome content. Incidence of stroke was monitored prospectively from the baseline examination in 1992 to 1994 until December 31, 2008. The case-finding procedures included a broad search among patients with neurological symptoms that could indicate stroke. Stroke was defined according to the WHO criteria. By definition, patients with transient ischemic attacks are excluded. The stroke subtypes are coded according to International Classification of Diseases revision 9. Cerebral infarction (International Classification of Diseases code 434) is diagnosed when CT, MRI, or autopsy verifies the infarction in location corresponding to the focal neurology or excludes hemorrhage and nonvascular disease. The ischemic strokes were retrospectively classified into etiological subtypes by review of hospital records. A board-certified neurologist with expertise in cerebrovascular diseases and a specialized research nurse reviewed the records. The TOAST^11^ and CCS^12^ criteria were applied.

**Osteoarthritis Initiative (OAI)**

The OAI is a publicly and privately funded prospective longitudinal cohort with a primary objective of identifying risk factors for incidence and progression of tibiofemoral knee osteoarthritis (OA). The OAI utilized a focused population-based recruitment to enroll 4,674 men and women between the ages of 45 – 79 years who either had radiographic symptomatic knee OA or who were without radiographic symptomatic OA in both knees but were considered high risk for OA because they had two or more known risk factors for knee OA. Subjects were recruited into the baseline phase of the OAI at multiple sites throughout the US between 2004 and 2006. All subjects were invited back for follow-up examinations to assess incidence or progression of OA annually, for up to 5 years. Phenotype data from the baseline and follow-up examinations are available for public access from the Osteoarthritis Initiative database. The Genetic Components of Knee Osteoarthritis (GeCKO) Study was initiated in 2009 as a genetic ancillary study to perform a genome-wide association study to identify genetic variants associated with radiographic osteoarthritis. This study included 4,482 individuals participating in the parent OAI study genotyped on the Illumina HumanOmni2.5M.

**Project MinE/Population based ALS registry, The Netherlands**

Dutch controls from cohorts NL4 (IlluminaOmniExpress) and NL5 (Illumina 2.5M) were included, originally described in Rheenen, W. van et al. Genome-wide association analyses identify new risk variants and the genetic architecture of amyotrophic lateral sclerosis. Nature Genetics 2016; 48, 1043–1048. The controls were identified through ongoing population based ALS registry, The Netherlands. (Huisman, M. H. B. et al. Population based epidemiology of amyotrophic lateral sclerosis using capture-recapture methodology. Journal of Neurology, Neurosurgery & Psychiatry 2011; 82, 1165–1170).

In brief, cases with ALS are diagnosed with probable or definite ALS according to the revised El-Escorial Criteria by neurologists specialized in motor neuron diseases. Both cases with and without a family history for ALS are included. Tertiary referral centers for ALS were University Medical Center Utrecht, Academic Medical Centre Amsterdam and Radboud University Medical Center Nijmegen. Both patients with and without a family history for ALS are included. Control individuals are free of any neuromuscular disease and matched for age, gender and ethnicity to cases.

2. **Genome-wide Association analysis**

Since many contributing study sites had small numbers of cases or may have lacked control samples, we consolidated datasets in analysis strata (Table S1) and matched cohorts based on genotype array, population structure, and whether or not they needed controls.^13^ We performed our association testing at the strata level and then used inverse variance meta-analysis (GWAMA) to combine results across strata using both fixed and random effects models.

Genotype data was filtered by individual missingness and SNP call rate. In addition, we removed snps that were out of Hardy-Weinberg equilibrium (p < 1x10^-5)^ in controls or if they had a minor allele frequency of less than 1%. Samples were removed if they had a sex-mismatch or had a high degree of relatedness to other samples.

Genotypes were lifted to build Hg38 if necessary and were imputed using the TOPMED imputation panel on the University of Michigan Imputation Server.

Two Step Meta-Analysis Overview

We first performed population-based meta-analysis for strata of recent African ancestry, European ancestry, and South Asian Ancestry. We required that each SNP be present in at least two analysis strata to be included in the ancestry-specific meta-analysis.

The second step was to perform a trans-ancestry meta-analysis by combining the ancestry-specific results into a single analysis. Following meta-analysis, we excluded SNPs with heterogenous effects across strata (post-meta i2 > 0.5) and those present in only a single stratum. For subtype-specific analysis, we also required a minor allele count (MAC) ≥ 30 within each stratum. Association analyses were performed using PLINK for European ancestry and SAIGE for Transethnic ancestry.

In order to compare our effect estimates in our early onset stroke GWAS, we performed a late age onset stroke analysis in cases over the age of 60 using the SiGN stroke dataset as described by Pulit et al.^13^ Logistic regression was performed using PLINK. Covariates in the logistic regression model included Age at first stroke or age of control at enrollment, sex, and the first 10 principal components to account for ancestry. Strata level results were combined using GWAMA package. Results were filtered by MAF ≥ 1% and R^2^ imputation quality score ≥ 0.5.

3. **Conditional Analysis at the *ABO* locus**

We performed a conditional analysis of stroke with other SNPs at the *ABO* locus after conditioning on associations of stroke with rs8176685 and rs529565. In addition to retaining these two SNPs as independent variables in a logistic regression analysis, we also included sex and up to 8 PCs as covariates. We tested the effects of 50 additional SNPs within 50 kb of the boundaries of the *ABO* gene. We flagged SNPs as showing evidence for residual association if the p-value for association was < 0.01. We restricted this analysis to samples of European ancestry because it is the largest population group within our cohort and has a more defined haplotype structure. As in the main GWAS, we performed the conditional analysis in PLINKv1.9 for each stratum and then combined results using meta-analysis.

4. **ABO blood group-determining SNPs, serologic ABO blood groups, and stroke**

4.1 Association of ABO haplotype-defining SNPs with stroke

Five SNPs tag define the 5 common ABO haplotypes as defined in Table below. We performed a genetic association analysis to determine associations of each blood group haplotype with early and late onset stroke. Analyses were performed within each analysis stratum, adjusting for age, sex and x PCs and results combined in meta-analysis.

| Table: 5 haplotypes that tag the major ABO blood groups* | | | | | | |  |  |
| --- | --- | --- | --- | --- | --- | --- | --- | --- |
| **ABO Blood group** | **rs2519093** | | **rs8176719** | **rs1053878** | **Rs8176743** | **rs41302905** | **Hapolotype freq** | **Effect on VTE** |
| O1 | C | | **delG** | G | C | C | 0.63 | ref |
| O2 | C | | G | G | C | **T** | 0.02 | ↓ |
| A1 | **T** | | G | G | C | C | 0.2 | ↑↑↑ |
| A2 | C | | G | **A** | C | C | 0.07 | ↑ |
| B | C | | G | G | **T** | C | 0.08 | ↑↑↑ |
| * adapted from Table 2 in Goumidi et al. (Blood 137:2394-2400, 2021)^14^ | | | | | | |  |  |

4.2 Distribution of ABO blood group among early onset stroke, late onset stroke, and controls

We defined serologic blood groups based on two SNPs rs8176746 and rs8176719, and rs5059223 as described in Groot et al.^15^ and then compared the distribution of ABO blood types among early and late onset stroke cases and controls. We initially tested for differences in the distribution of ABO blood groups between early vs late onset cases, early cases vs controls, and late cases vs controls using a 3 df chi-square omnibus (4 blood groups by 2 comparison groups). If an overall difference was observed between groups, we then performed pairwise tests to identify which blood groups differed significantly between the two comparison groups (e.g., distribution of blood group A vs non-A between early and late).

Genetically Determined Serologic Blood Group Coding Scheme (from Groot, 2020)

| **rsID** | **Alleles** |  |  |  |  |  |  |  |
| --- | --- | --- | --- | --- | --- | --- | --- | --- |
| rs8176719 | TC_TC(/T) | 0 | 0 | 1 | 2 | 2 | 1 | 2 |
| rs8176746 | G:T_T(/G) | 0 | 1 | 0 | 0 | 2 | 1 | 1 |
|  | **Serologic Blood Group Assignment** | **O** | **O** | **A** | **A** | **B** | **B** | **AB** |

5. **Association of *ABO* SNPs with early and late onset stroke in UKB**

Ischemic stroke cases were extracted by ICD code algorithm previously published in the “Definitions of Stroke for UK Biobank Phase 1 Outcomes Adjudication” (see <https://biobank.ndph.ox.ac.uk/showcase/label.cgi?id=43>) and validated by stroke physician review of electronic patient records.^16^ Controls were matched to cases on age, sex, and ancestry, with up to 20 controls chosen per case. Any individual related to an already selected individual was removed. Age at Ischemic Stroke diagnosis was determined at either the main or secondary diagnosis, whichever was younger. We defined early onset Ischemic Stroke cases as subjects with age at diagnosis before age 60 and late onset Ischemic Stroke cases as those with age at diagnosis at age 60 or older. Our analysis included 1,803 early onset cases and 4,071 late onset cases.

PLINK was used to perform logistic regression using age, sex and 5 principal components for ancestry.

ICD10 Codes used to define Ischemic Stroke

ICD 10 Code I63 Cerebral infarction

ICD 10 Code I63.0 Cerebral infarction due to thrombosis of precerebral arteries

ICD 10 Code I63.1 Cerebral infarction due to embolism of precerebral arteries

ICD 10 Code I63.2 Cerebral infarction due to unspecified occlusion or stenosis of precerebral arteries

ICD 10 Code I63.3 Cerebral infarction due to thrombosis of cerebral arteries

ICD 10 Code I63.4 Cerebral infarction due to embolism of cerebral arteries

ICD 10 Code I63.5 Cerebral infarction due to unspecified occlusion or stenosis of cerebral arteries

ICD 10 Code I63.6 Cerebral infarction due to cerebral venous thrombosis, nonpyogenic

ICD 10 Code I63.8 Other cerebral infarction

ICD 10 Code I63.9 Cerebral infarction, unspecified

ICD 10 Code I64 Stroke, not specified as haemorrhage or infarction

Source: Definitions of Stroke for UK Biobank Phase 1 Outcomes Adjudication. Version 1.1. August 2017

6. **Association of ABO SNPs with early and late onset VTE in UKB**

Individuals were defined as a VTE case based on at least one of the following ICD-10 codes being recorded in their medical record as either a main or secondary diagnosis:

1. Hospitalization for ICD-10 Code I80.1 or I80.2 - phlebitis and thrombophlebitis of the femoral vein or other deep vessels of lower extremities;
2. Hospitalization for ICD-10 Code I82.2 - embolism and thrombosis of vena cava;
3. Hospitalization for ICD-10 Code I26.0 or I26.9 - pulmonary embolism with or without acute cor pulmonale;

Controls were selected after exclusion of related codes. Controls were matched to cases on age, sex, and ancestry, with up to 5 controls chosen per case. Any individual related to an already selected individual was removed. Age at VTE diagnosis was determined at either the main or secondary diagnosis, whichever was younger. We defined early onset VTE cases as subjects with age at diagnosis before age 60 and late onset VTE cases as those with age at diagnosis at age 60 or older. Our analysis included 3,519 early onset cases and 5,046 late onset cases.

7. **Association of VTE PRS with early and late onset stroke**

Additive, effect weighted polygenic risk scores were calculated for VTE based on a published list of 297 VTE risk alleles identified through a large primary GWAS (Klarin et al, 2019).^17^ This prior VTE PRS was constructed using pruning and thresholding criteria of R2 < 0.2 and P < 1 × 10^−5^) and obtained from European MVP v.2.1 and UK Biobank European VTE meta-analyzed summary statistics. Of these 297 SNPs, 255 were available across all strata of our early and late onset GWAS as used for analysis. We tested the association between the VTE PRS score with stroke in the European ancestry sample using logistic regression with 10 PCs and sex included as covariates. Scores were generated using PRSice^18^ and the -score avg option. Association analysis with stroke was performed in RStudio. We computed odds ratios corresponding to the associations of VTE PRS on early and late onset stroke and tested for homogeneity of the odds ratios using the Wald test.

8. **Co-location of associations at the ABO locus between EOS and other thrombotic-related disorders and biomarkers**

We performed colocalization analyses to evaluate evidence for a shared common variant at the ABO locus between early onset stroke and late onset stroke and between early onset stroke and 3 prothrombotic traits (VTE and plasma levels of von Willebrand factor (VWF) and Factor VIII (FVIII). For this analysis, we considered all SNPs with 50 kb of the gene boundaries. Analyses were performed using summary genetic association results with the coloc software program.^19^ Summary statistics for GWAS results were obtained from the sources indicated in the table below. Briefly, coloc utilizes a Bayesian approach and summary level association level results for two traits to calculate the posterior probabilities of five competing models that assess whether the associations are due to the same or a different causal variant. The competing models tested are:

H_0_: no causal SNP for either trait

H_1_: causal SNP for Trait 1, not Trait 2

H_2_: causal SNP for Trait 2, not Trait 1

H_3_: different causal SNPs for Traits 1 and 2

H_4_: shared causal SNP for Traits 1 and 2

For each set of traits, we considered a posterior probability >80% for H4 as indicating strong evidence for a shared causal variant for both traits.

| **Disorder/Trait** | **Source of summary statistics** | **Sample size** | **Source/Publication** |
| --- | --- | --- | --- |
| Late onset stroke | GWAS of late onset stroke from SiGN | 9,272 cases  25,124 controls | Described in this manuscript |
| VTE | UKB | 4,620 cases  356,574 controls | Internal analysis; ICD codes |
| Plasma VWF levels | CHARGE Consortium | 46,354 subjects | Sabater-Lleal M, Huffman JE, de Vries PS, et al. Genome-Wide Association Transethnic Meta-Analyses Identifies Novel Associations Regulating Coagulation Factor VIII and von Willebrand Factor Plasma Levels. Circulation. 2019;139(5):620-635. |
| Plasma Factor VIII levels | CHARGE Consortium | 46,354 subjects | Sabater-Lleal M, et al., Circulation. 2019;139(5):620-635. |
