## Appendix for "Genetic Contributions to Early and Late Onset Ischemic Stroke"

**eAppendix1**

1. **CADISP CONSORTIUM INVESTIGATORS**

**CADISP Consortium**

Shérine Abboud^1^, Margareth Amort^2^, Marie-Luise Arnold^3^, Yannick Béjot,^4^ Simone Beretta^5^, Anna Bersano^6^, Marie Bodenant^7^, Leo Bonati^2^, Tobias Brandt^8^, Sandrine Canaple^9^, Filomena Caria^10^, John Cole^11^, Sophie Crozier^12^, Jean Dallongeville^13^, Valeria De Giuli^10^, Stéphanie Debette^14^, Pierre Decavel^15^, Sandrine Deltour^12^, Martin Dichgans^16^, Michael Dos Santos^17^, Stefan Engelter^2^, Carlo Ferrarese^5^, Felix Fluri^2^, Henrik Gensicke^2^, Giacomo Giacolone^18^, Maurice Giroud^4^, Dominique Gisler^2^, Olivier Godefroy^9^, Armin Grau^17^, Caspar Grond-Ginsbach^3^, Florian Hatz^2^, Steven Kittner^11^, Manja Kloss^3^, Silvia Lanfranconi^6^, Sara Leder^12^, Anne Léger^12^, Didier Leys^7^, Christoph Lichy^3^, Fabien Louillet^19^, Philippe Lyrer^2^, Jennifer Majersik^20^, Hugh Markus^21^, Jean-Louis Mas^19^, Elizabeth Medeiros^15^, Isabelle Méresse^12^, Antti J Metso^22^, Tiina Metso^22^, Braxton Mitchell^11^, Paola Montiel^15^, Thierry Moulin^15^, Alessandro Padovani^10^, Massimo Pandolfo^1^, Stefano Paolucci^23^, Alessandro Pezzini^10^, Loris Poli^10^, Yves Samson^12^, Andrew Southerland^24^, Turgut Tatlisumak^22^, Vincent Thijs^25^, Constanze Thomas-Feles^8^, Emmanuel Touzé^19^, Christopher Traenka^2^, Fabrice Vuillier^15^, Ralf Weber^8^, Inge Werner^3^, Bradford Worrall^24^

**Affiliations**

^1^ Departments of Neurology, Erasmus University Hospital, Brussels and Laboratory of Experimental Neurology, ULB, Brussels

^2^ Department of Neurology, Basel University Hospital

^3^ Departments of Neurology, Heidelberg University Hospital

^4^ Dijon University Hospital

^5^  University of Milano Bicocca, San Gerardo Hospital, Monza, Italy

^6^ Milan University Hospital

^7^ Departments of Neurology, Lille University Hospital-Inserm U1171

^8^ Department of Rehabilitation: Schmieder-Klinik, Heidelberg

^9^ Amiens University Hospital

^10^ Departments of Neurology: Brescia University Hospital

^11^ Department of Neurology, University of Maryland, Baltimore, USA

^12^ Pitié-Salpêtrière University Hospital, Paris

^13^ Inserm U1167, Pasteur Institute, Lille

^14^ Department of Neurology Bordeaux University Hospital and Inserm U1219 University of Bordeaux

^15^ Besançon University Hospital

^16^  University Hospital of München

^17^ University Hospital of Ludwigshafen

^18^ Milan Scientific Institute San Raffaele University Hospital

^19^ Sainte-Anne University Hospital, Paris

^20^ Department of Neurology, University of Utah, Salt Lake City, USA

^21^ Clinical Neuroscience, Cambridge University

^22^ Department of Neurology, Helsinki University Central Hospital, Helsinki

^23^ Department of Rehabilitation, Santa Lucia Hospital, Rome

^24^ Department of Neurology, University of Virginia, Charlottesville, USA

^25^ Department of Neurology, Leuven University Hospital

2. **INVENT CONSORTIUM INVESTIGATORS**

Philippe Amouyel,^1^ Mariza de Andrade,^2^ Saonli Basu,^3^ Claudine Berr,^4^ Jennifer A Brody,^5^ Daniel I Chasman,^6^ Jean-Francois Dartigues,^7^ Aaron R Folsom^,8^ Marine Germain,^9^ Hugoline de Haan,^10^ John Heit,^11^ Jeanine Houwing-Duitermaat,^12^ Christopher Kabrhel,^13^ Peter Kraft,^14^ Grégoire Legal,^15,16^ Sara Lindström,^14^ Ramin Monajemi,^12^ Pierre-Emmanuel Morange,^17^ Bruce M Psaty,^5,18^ Pieter H Reitsma,^19^ Paul M Ridker^,20^ Lynda M Rose,^21^ Frits R Rosendaal,^10^ Noémie Saut,^17^ Eline Slagboom,^22^ , David Smadja^23^ Nicholas L Smith,^18,24,25^ Pierre Suchon,^17^ Weihong Tang,^8^ Kent D Taylor,^26^ David-Alexandre Trégouët,^9^ Christophe Tzourio,^27^ Marieke CH de Visser,^19^ Astrid van Hylckama Vlieg,^10^ Lu-Chen Weng,^8^ Kerri L Wiggins,^24^

**Affiliations**

^1^ Institut Pasteur de Lille, Université de Lille Nord de France, INSERM UMR_S 744, Lille, France; Centre Hospitalier Régional Universitaire de Lille, Lille, France.

^2^ Division of Biomedical Statistics and Informatics Mayo Clinic, Rochester, MN, USA.

^3^ University of Minnesota, Division of Biostatistics, Minneapolis, MN, USA.

^4^ INSERM Research Unit U1061 , University of Montpellier I , Montpellier, France.

^5^ Cardiovascular Health Research Unit, Departments of Medicine, Epidemiology, and Health Services, University of Washington, Seattle, WA, USA.

^6^ Division of Preventive Medicine, Brigham and Women’s Hospital and Harvard Medical School, Boston, MA 02215, USA.

^7^ INSERM Research Center U897, University of Bordeaux, Bordeaux, France.

^8^ University of Minnesota, Division of Epidemiology and Community Health Minneapolis, MN, USA.

^9^ Institut National pour la Santé et la Recherche Médicale (INSERM), Unité Mixte de Recherche en Santé (UMR_S) 1166, F-75013, Paris, France; Sorbonne Universités, Université Pierre et Marie Curie (UPMC Univ Paris 06), UMR_S 1166, Team Genomics & Pathophysiology of Cardiovascular Diseases, F-75013, Paris, France; Institute for Cardiometabolism and Nutrition (ICAN), F-75013, Paris, France.

^10^ Department of Thrombosis and Hemostasis, Leiden University Medical Center, Leiden, The Netherlands; Department of Clinical Epidemiology, Leiden University Medical Center, Leiden, The Netherlands.

^11^ Division of Cardiovascular Diseases, Mayo Clinic, Rochester, MN, USA.

^12^ Department of Medical Statistics and Bioinformatics, Leiden University Medical Center, 2300 RC Leiden, Netherlands.

^13^ Department of Emergency Medicine, Massachusetts General Hospital, Channing Network Medicine, Harvard Medical School, Boston, MA 2114, USA.

^14^ Program in Genetic Epidemiology and Statistical Genetics, Department of Epidemiology, Harvard School of Public Health, Boston, MA 2115, USA.

^15^ Université de Brest, EA3878 and CIC1412, Brest, France.

^16^ Ottawa Hospital Research Institute at the University of Ottawa, Ottawa, ON, Canada.

^17^ Laboratory of Haematology, La Timone Hospital, F-13385, Marseille, France; INSERM, UMR_S 1062, Nutrition Obesity and Risk of Thrombosis,F-13385, Marseille, France; AixMarseille University, UMR_S 1062, Nutrition Obesity and Risk of Thrombosis, F-13385, Marseille, France.

^18^ Group Health Research Institute, Group Health Cooperative, Seattle WA 98101, USA.

^19^ Einthoven Laboratory for Experimental Vascular Medicine, Department of Thrombosis and Hemostasis, Leiden University Medical Center, Leiden,2300 RC, Netherlands.

^20^ Division of Preventive Medicine, Brigham and Women’s Hospital and Harvard Medical School, Boston, MA 02215, USA.

^21^ Division of Preventive Medicine, Brigham and Women’s Hospital, Boston, MA 02215, USA.

^22^ Department of Molecular Epidemiology, Leiden University Medical Center, 2300 RC Leiden, Netherlands.

^23^ Université Paris Descartes, Sorbonne Paris Cité, Paris, France; AP-HP, Hopital Européen Georges Pompidou, Service d’hématologie Biologique, Paris, France; INSERM, UMR_S 1140, Faculté de Pharmacie, Paris, France.

^24^ Department of Epidemiology, University of Washington, Seattle WA 98195, USA.

^25^ Seattle Epidemiologic Research and Information Center, VA Office of Research and Development, Seattle WA 98108, USA.

^26^ Los Angeles Biomedical Research Institute and Department of Pediatrics, Harbor-UCLA Medical Center, Torrence CA 90502, USA.

27 INSERM Research Center U897, University of Bordeaux, Bordeaux, France.
3. **Funding and Acknowledgements**

**Case-only & Case and Control Cohort:**

**ASGC:** Australian population control data were derived from the Hunter Community Study. We also thank the University of Newcastle for funding and the men and women of the Hunter region who participated in this study. This research was funded by grants from the Australian National and Medical Health Research Council (NHMRC Project Grant ID: 569257), the Australian National Heart Foundation (NHF Project Grant ID: G 04S 1623), the University of Newcastle, the Gladys M Brawn Fellowship scheme, and the Vincent Fairfax Family Foundation in Australia. Elizabeth G Holliday was supported by a Fellowship from the National Heart Foundation and National Stroke Foundation of Australia (ID: 100071).

**BASICMAR:** The Base de Datos de Ictus del Hospital del Mar (BASICMAR) Genetic Study was supported by the Ministerio de Sanidad y Consumo de España, Instituto de Salud Carlos III (ISC III) with the grants: Registro BASICMAR Funding for Research in Health (PI051737); GWA Study of Leukoaraiosis (GWALA) project from Fondos de Investigación Sanitaria ISC III (PI10/02064) and (PI12/01238); Agència de Gestió Ajuts Universitaris de Recerca (2014 SGR 1213) and Fondos European Regional Development Funding (FEDER/EDRF) Red INVICTUS-PLUS (RD16/0019/0002). Additional support was provided by the Fundació la Marató TV3 with the grant GODS project. Genestroke Consortium (76/C/2011) Recercaixa’13 (JJ086116). Assistance with data cleaning was provided by the Research in Cardiovascular and Inflammatory Diseases Program of Institute Hospital del Mar of Medical Investigations, Hospital del Mar, and the Barcelona Biomedical Research Park.

**Biobank Japan:** We would like to express our gratefulness to all the members of J-MICC, JPHC, and TMM. We extend our appreciation to staffs of BBJ for their outstanding assistance. This study was funded by the BioBank Japan project, which is supported by the Ministry of Education, Culture, Sports, Sciences and Technology (MEXT) of Japanese government and the Japan Agency for Medical Research and Development (AMED).

**BRAINS:** Biorepository of DNA in Stroke (BRAINS) is partly funded by a Senior Fellowship from the Department of Health (UK) to P Sharma, the Henry Smith Charity and the UK-India Education Research Institutive (UKIERI) from the British Council.

**CADISP:** The Cervical Artery Dissections and Ischemic Stroke Patients (CADISP) study has been supported by Inserm, Lille 2 University, Institut Pasteur de Lille and Lille University Hospital and received funding from the ERDF (FEDER funds) and Région Nord-Pas de Calais in the frame of Contrat de Projets Etat-Region 2007-2013 Région Nord-Pas-de-Calais - Grant N°09120030, Centre National de Genotypage, Emil Aaltonen Foundation, Paavo Ilmari Ahvenainen Foundation, Helsinki University Central Hospital Research Fund, Helsinki University Medical Foundation, Päivikki and Sakari Sohlberg Foundation, Aarne Koskelo Foundation, Maire Taponen Foundation, Aarne and Aili Turunen Foundation, Lilly Foundation, Alfred Kordelin Foundation, Finnish Medical Foundation, Orion Farmos Research Foundation, Maud Kuistila Foundation, the Finnish Brain Foundation, Biomedicum Helsinki Foundation, Projet Hospitalier de Recherche Clinique Régional, Fondation de France, Génopôle de Lille, Adrinord, Basel Stroke-Funds, Käthe-Zingg-Schwichtenberg-Fonds of the Swiss Academy of Medical Sciences, Swiss Heart Foundation.

**China Kadoorie Biobank:** CKB acknowledges the contribution of participants, project staff, and the China National Centre for Disease Control and Prevention (CDC) and its regional offices. China Kadoorie Biobank was supported as follows: Baseline survey and first re-survey: Hong Kong Kadoorie Charitable Foundation; long-term follow-up and second re-survey: UK Wellcome Trust (212946/Z/18/Z, 202922/Z/16/Z, 104085/Z/14/Z, 088158/Z/09/Z), National Natural Science Foundation of China (91843302), and National Key Research and Development Program of China (2016YFC 0900500, 0900501, 0900504, 1303904). DNA extraction and genotyping: GlaxoSmithKline, UK Medical Research Council (MC_PC_13049, MC-PC-14135). The UK Medical Research Council (MC_UU_00017/1, MC_UU_12026/2 MC_U137686851), Cancer Research UK (C16077/A29186; C500/A16896), and the British Heart Foundation (CH/1996001/9454) provide core funding to the Clinical Trial Service Unit and Epidemiological Studies Unit at Oxford University for the project.

**Edinburgh:** The Edinburgh Stroke Study was supported by the Wellcome Trust and the Binks Trust. Sample processing occurred in the Genetics Core Laboratory of the Wellcome Trust Clinical Research Facility, Western General Hospital, Edinburgh, UK. Dr. Rannikmäe was funded from HDR UK fellowship MR/S004130/1

**EPIC-CVD:** This work was supported by core funding from the: UK Medical Research Council (MR/L003120/1), British Heart Foundation (RG/13/13/30194; RG/18/13/33946), BHF Centre for Research Excellence (RE/18/1/34212) and NIHR Cambridge Biomedical Research Centre (BRC-1215-20014) [*]. EPIC-CVD was also funded by the European Research Council (268834) and the European Commission Framework Programme 7 (HEALTH-F2-2012-279233).

*The views expressed are those of the author(s) and not necessarily those of the NIHR or the Department of Health and Social Care.

The coordination of EPIC is financially supported by International Agency for Research on Cancer (IARC) and also by the Department of Epidemiology and Biostatistics, School of Public Health, Imperial College London which has additional infrastructure support provided by the NIHR Imperial Biomedical Research Centre (BRC). The national cohorts are supported by: Danish Cancer Society (Denmark); Ligue Contre le Cancer, Institut Gustave Roussy, Mutuelle Générale de l’Education Nationale, Institut National de la Santé et de la Recherche Médicale (INSERM) (France); German Cancer Aid, German Cancer Research Center (DKFZ), German Institute of Human Nutrition PotsdamRehbruecke (DIfE), Federal Ministry of Education and Research (BMBF) (Germany); Associazione Italiana per la Ricerca sul Cancro-AIRC-Italy, Compagnia di SanPaolo and National Research Council (Italy); Dutch Ministry of Public Health, Welfare and Sports (VWS), Netherlands Cancer Registry (NKR), LK Research Funds, Dutch Prevention Funds, Dutch ZON (Zorg Onderzoek Nederland), World Cancer Research Fund (WCRF), Statistics Netherlands (The Netherlands); Health Research Fund (FIS) - Instituto de Salud Carlos III (ISCIII), Regional Governments of Andalucía, Asturias, Basque Country, Murcia and Navarra, and the Catalan Institute of Oncology - ICO (Spain); Swedish Cancer Society, Swedish Research Council and County Councils of Skåne and Västerbotten (Sweden); Cancer Research UK (14136 to EPIC-Norfolk; C8221/A29017 to EPIC-Oxford), Medical Research Council (1000143 to EPIC-Norfolk; MR/M012190/1 to EPIC-Oxford). (United Kingdom).

We thank all EPIC participants and staff for their contribution to the study, the laboratory teams at the Medical Research Council Epidemiology Unit for sample management and Cambridge Genomic Services for genotyping, Sarah Spackman for data management, and the team at the EPIC-CVD Coordinating Centre for study coordination and administration.

**FUTURE/Odyssey:** F-E d L received a grant from the Dutch Heart Foundation (grant 2014 T060 to FE.d.L) and Bike4Brains. AMT received a grant from the Dutch Heart Foundation (grant 2016 T044 to A.M.T) and from the Netherlands CardioVascular Research Initiative: the Dutch Heart Foundation (CVON 2018-28 & 2012-06 Heart Brain Connection to A.M.T)

**MGH-GASROS:** MGH Genes Affecting Stroke Risk and Outcome Study (MGH-GASROS) was supported by NINDS (U01 NS069208), the American Heart Association/Bugher Foundation Centers for Stroke Prevention Research 0775010N, the NIH and NHLBI's STAMPEED genomics research program (R01 HL087676), and a grant from the National Center for Research Resources. The Broad Institute Center for Genotyping and Analysis is supported by grant U54 RR020278 from the National Center for Research resources.

**GCNKSS:** Greater Cincinnati/Northern Kentucky Stroke Study (GCNKSS) was supported by the NIH (NS030678).

**Geisinger:** Dr Abedi has research support from the Defense Threat Reduction Agency (DTRA) grant No. HDTRA1-18-1-0008 and from National Institutes of Health grant No. R56HL116832. Dr Zand has research support from Bucknell University Initiative Program, Roche—Genentech Biotechnology Company, and the Geisinger Health Plan Quality fund.

**GEOS:** Genetics of Early Onset Stroke (GEOS) Study, Baltimore, USA was supported by the NIH Genes, Environment and Health Initiative (GEI) Grant U01 HG004436, as part of the GENEVA consortium under GEI, with additional support provided by the Mid-Atlantic Nutrition and Obesity Research Center (P30 DK072488), and the Office of Research and Development, Medical Research Service, and the Baltimore Geriatrics Research, Education, and Clinical Center of the Department of Veterans Affairs. Genotyping services were provided by the Johns Hopkins University Center for Inherited Disease Research (CIDR), which is fully funded through a federal contract from the NIH to the Johns Hopkins University (contract number HHSN268200782096C). Assistance with data cleaning was provided by the GENEVA Coordinating Center (U01 HG 004446; PI Bruce S Weir). Study recruitment and assembly of datasets were supported by a Cooperative Agreement with the Division of Adult and Community Health, Centers for Disease Control and Prevention and by grants from NINDS and the NIH Office of Research on Women's Health (R01 NS45012, U01 NS069208- 01).

**GRAZ:** The Austrian Stroke Prevention Study was supported by the Austrian Science Fund (FWF) grant Nos. P20545-P05 and P13180 and I904-B13 (Era-Net). The Medical University of Graz supports the databases of the Graz Stroke Study and the Austrian Stroke Prevention Study.

**Helsinki:** The study was supported by the Finnish Medical Foundation, Sigrid Juselius Foundation, and the Helsinki University Central Hospital governmental subsidiary funds for clinical research. The investigators thank Marja Metso, RN for her support.

**INTERSTROKE:** The INTERSTROKE study was supported by the Canadian Institutes of Health Research, Heart and Stroke Foundation of Canada, Canadian Stroke Network, Health Research Board Ireland, Swedish Research Council, Swedish Heart and Lung Foundation, The Health & Medical Care Committee of the Regional Executive Board, Region Västra Götaland (Sweden), AstraZeneca, Boehringer Ingelheim (Canada), Pfizer (Canada), MSD, Chest, Heart and Stroke Scotland, and The Stroke Association, with support from The UK Stroke Research Network. Microarray genotyping for a subset of INTERSTROKE participants was funded by the Heart and Stroke Foundation of Canada Grant NA‐6872 (PI: Drs Pare, Anand, O’Donnell, Xie, Yusuf).

**INVENT:** David-Alexandre Trégouët is supported by the «EPIDEMIOM-VTE» Senior Chair from the Initiative of Excellence of the University of Bordeaux.

**ISGS:** The Ischemic Stroke Genetics Study (ISGS) was supported by the NINDS (R01 NS42733; PI Dr Meschia). Both SWISS and ISGS received additional support, in part, from the Intramural Research Program of the National Institute on Aging (Z01 AG000954-06; PI Andrew Singleton). SWISS and ISGS used samples and clinical data from the NIH-NINDS Human Genetics Resource Center DNA and Cell Line Repository (http://ccr.coriell.org/ninds), human subject protocol Nos. 2003-081 and 2004-147. SWISS and ISGS used stroke-free participants from the Baltimore Longitudinal Study of Aging (BLSA) as controls with the permission of Dr Luigi Ferrucci. The inclusion of BLSA samples was supported, in part, by the Intramural Research Program of the National Institute on Aging (Z01 AG000015-50), human subject protocol No. 2003-078. This study used the high-performance computational capabilities of the Biowulf Linux cluster at the NIH (http://biowulf.nih.gov). For SWISS and ISGS cases of African ancestry, a subset of the Healthy Aging in Neighborhoods of Diversity across the Life Span study (HANDLS) were used as stroke-free controls. HANDLS is funded by the National Institute of Aging (1Z01AG000513; PI Michele K. Evans).

**Krakow:** Phenotypic data and genetic specimens collection were funded by the grant from the Polish Ministry of Science and Higher Education for Leading National Research Centers (KNOW) and by the grants from the Jagiellonian University Medical College in Krakow, Poland: K/ZDS/002848, K/ZDS/003844.

**Leuven:** The Leuven Stroke genetics study was supported by personal research funds from the Department of Neurology of the University Hospitals Leuven. Dr. Lemmens is a Senior Clinical Investigator of FWO Flanders (FWO 1841918N).

**Lund:** Supported by the Swedish Research Council (2019-01757), The Swedish Heart-Lung Foundation, Region Skåne, Skåne University Hospital, the Freemasons Lodge of Instruction Eos in Lund, King Gustaf V’s and Queen Victoria’s Foundation, Lund University, CaNVAS NIH (1R01NS114045-01), and The Swedish Government (under the “Avtal om Läkarutbildning och Medicinsk Forskning, ALF”). Biobank services were provided by Region Skåne Competence Centre (RSKC Malmö), Skåne University Hospital, Malmö, Sweden, and Biobank, Labmedicin Skåne, University and Regional Laboratories Region Skåne, Sweden.

**MCISS:** The Middlesex County Ischemic Stroke Study (MCISS) was supported by intramural funding from the New Jersey Neuroscience Institute/JFK Medical Center, Edison, NJ, and The Neurogenetics Foundation, Cranbury, NJ. We acknowledge Dr Souvik Sen for his advice and encouragement in the initiation and design of this study.

**MIAMISR:** The Cerebrovascular Biorepository at University of Miami/Jackson Memorial Hospital (The Miami Stroke Registry, Institutional Review Board No. 20070386) was supported by the Department of Neurology at University of Miami Miller School of Medicine and Evelyn McKnight Brain Institute. Biorepository and DNA extraction services were provided by the Hussmann Institute for Human Genomics at the Miller School of Medicine.

**Milano:** Milano - Besta Stroke Register Collection and genotyping of the Milan cases within CEDIR were supported by the Italian Ministry of Health (Grant Numbers: RC 2007/LR6, RC 2008/LR6; RC 2009/LR8; RC 2010/LR8; GR-2011-02347041). FP6 LSHM-CT-2007-037273 for the PROCARDIS control samples.

**Munich:** This project received funding from the European Union’s Horizon 2020 research and innovation programme (666881), SVDs@target (to MD; 667375)); the DFG as part of the Munich Cluster for Systems Neurology (EXC 2145 SyNergy – ID 390857198), the CRC 1123 (B3; to MD), DI 722/16-1 (project ID: 428668490), DI 722/13-1; and the Fondation Leducq (Transatlantic Network of Excellence on the Pathogenesis of Small Vessel Disease of the Brain; to MD).

Institute for Stroke and Dementia Research (ISD), University Hospital, LMU Munich, Munich, Germany. Munich Cluster for Systems Neurology (SyNergy), Munich, Germany. German Center for Neurodegenerative Diseases (DZNE), Munich, Germany.

**NHS:** The Nurses’ Health Study work on stroke is supported by grants from the NIH, including HL088521 and HL34594 from the National Heart, Lung, and Blood Institute, as well as grants from the National Cancer Institute funding the questionnaire follow-up and blood collection: CA87969 and CA49449.

**NOMAS:** The Northern Manhattan Study (NOMAS) was supported by grants from the NINDS (R01 NS029993, R01 NS27517). The Cerebrovascular Biorepository at University of Miami/Jackson Memorial Hospital (The Miami Stroke Registry, Institutional Review Board No. 20070386) was supported by the Department of Neurology at University of Miami Miller School of Medicine and Evelyn McKnight Brain Institute. Biorepository and DNA extraction services were provided by the Hussmann Institute for Human Genomics at the Miller School of Medicine.

**OXVASC:** The Oxford Vascular Study was supported by the Wellcome Trust, Wolfson Foundation, Stroke Association, Medical Research Council, Dunhill Medical Trust, NIH Research (NIHR), and NIHR Oxford Biomedical Research Centre based at Oxford University Hospitals NHS Trust and University of Oxford. Dr Rothwell is in receipt of Senior Investigator Awards from the Wellcome Trust and the NIHR.

**RACE:** We are thankful to the RACE study participants. Fieldwork in RACE was funded by the R-21 grant provided by the NINDS and the Fogarty International Center (1R21NS064908-01) and educational grants available to Dr. Saleheen at the Center for Non-Communicable Diseases, Pakistan. We would also like to acknowledge the contributions made by Professor John Danesh, Dr. Ayeesha Kamal and Professor Panos Deloukas.

**REGARDS:** The Reasons for Geographic and Racial Differences in Stroke (REGARDS) Study was supported by a cooperative agreement U01 NS041588 from the NINDS, NIH, and Department of Health and Human Service. A full list of participating REGARDS investigators and institutions can be found at <http://www.regardsstudy.org>.

**SAHLSIS:** The Sahlgrenska Academy Study on Ischemic Stroke was supported by the Swedish Research Council (2018-02543), the Swedish Heart and Lung Foundation (20190203), the Swedish state under the agreement between the Swedish government and the county councils, the ALF-agreement (ALFGBG-720081).

**SIFAP:**  The SIFAP study (Stroke in Young Fabry Patients, http://www.sifap.eu; ClinicalTrials.gov: NCT00414583) has been supported partially by an unrestricted scientific grant from Shire Human Genetic Therapies. Funding for genotyping and analysis of samples were supported by the National Institutes of Health Genes, Environment and Health Initiative (GEI) Grant U01 HG004436, as part of the GENEVA consortium.

**SLESS:** This work was supported by a Stroke Association (UK) Programme Grant (PROG 3), the National Institute for Health Research Biomedical Research Centre (NIHR BRC) at South London and Maudsley NHS Foundation Trust and the National Institute for Health Research (NIHR) Biomedical Research Centre based at Guy's and St Thomas' NHS Foundation Trust and King's College London.

S.Bell and H.S.M. are funded by the British Heart Foundation (RG/16/4/32218). H.S.M. is also supported by an NIHR Senior Investigator award. This research was supported by the NIHR Cambridge Biomedical Research Centre (BRC-1215-20014).  The views expressed are those of the author(s) and not necessarily those of the NIHR or the Department of Health and Social Care

**SPS3:** The Secondary Prevention of Small Subcortical Strokes trial was funded by the US National Institute of Health and Neurological Disorders and Stroke grant No. U01NS38529-04A1 (principal investigator, Oscar R. Benavente; coprincipal investigator, Robert G. Hart). The SPS3 Genetic Substudy (SPS3-GENES) was funded by R01 NS073346 (coprincipal investigators, Julie A. Johnson, Oscar R. Benavente, and Alan R. Shuldiner) and U01 GM074492-05S109 (principal investigator, Julie A. Johnson).

**St. George:** The principal funding for this study was provided by the Wellcome Trust, as part of the Wellcome Trust Case Control Consortium 2 project (085475/B/08/Z and 085475/Z/08/Z and WT084724MA). Collection of some of the St George’s stroke cohort was supported by project grant support from the Stroke Association.

**SWISS:** The Sibling with Ischemic Stroke Study (SWISS) was supported by the NINDS (R01 NS39987; PI Dr Meschia). Both SWISS and ISGS received additional support, in part, from the Intramural Research Program of the National Institute on Aging (Z01 AG000954-06; PI Andrew Singleton). SWISS and ISGS used samples and clinical data from the NIH-NINDS Human Genetics Resource Center DNA and Cell Line Repository (http://ccr.coriell.org/ninds), human subject protocol Nos. 2003-081 and 2004-147. SWISS and ISGS used stroke-free participants from the Baltimore Longitudinal Study of Aging (BLSA) as controls with the permission of Dr Luigi Ferrucci. The inclusion of BLSA samples was supported, in part, by the Intramural Research Program of the National Institute on Aging (Z01 AG000015-50), human subject protocol No. 2003-078. This study used the high-performance computational capabilities of the Biowulf Linux cluster at the NIH (http://biowulf.nih.gov). For SWISS and ISGS cases of African ancestry, a subset of the Healthy Aging in Neighborhoods of Diversity across the Life Span study (HANDLS) were used as stroke-free controls. HANDLS is funded by the National Institute of Aging (1Z01AG000513; PI Michele K. Evans).

**UK Biobank:** UK Biobank has received funding from the UK Medical Research Council, Wellcome Trust, Department of Health, British Heart Foundation, Diabetes UK, Northwest Regional Development Agency, Scottish Government, and Welsh Assembly Government.

**VHIR-FMT-Barcelona:** The Barcelona GWAs Study was supported by the Genetic contribution to functional Outcome and Disability after Stroke (GODS) project and EPIGENESIS project Fundació la Marató de TV3, GENERACION Project (Instituto de Salud Carlos III: PI15/01978), Maestro Project (Instituto de Salud Carlos III,  Fondo Europeo de Desarrollo Regional (FEDER)).) and by the Miguel Servet grant (Pharmastroke project: CP12/03298). I. F-C. is supported by the Miguel Servet programme (CP12/03298), Instituto de Salud Carlos III.

**VISP:**  The GWAS component of the VISP study was supported by the United States National Human Genome Research Institute (NHGRI), Grant U01 HG005160 (PI Michèle Sale & Bradford Worrall), as part of the Genomics and Randomized Trials Network (GARNET). Genotyping services were provided by the Johns Hopkins University Center for Inherited Disease Research (CIDR), which is fully funded through a federal contract from the NIH to the Johns Hopkins University. Assistance with data cleaning was provided by the GARNET Coordinating Center (U01 HG005157; PI Bruce S Weir). Study recruitment and collection of datasets for the VISP clinical trial were supported by an investigator-initiated research grant (R01 NS34447; PI James Toole) from the United States Public Health Service, NINDS, Bethesda, Maryland. Control data for comparison with European ancestry VISP stroke cases were obtained through the database of genotypes and phenotypes (dbGAP) High Density SNP Association Analysis of Melanoma: Case-Control and Outcomes Investigation (phs000187.v1.p1; R01CA100264, 3P50CA093459, 5P50CA097007, 5R01ES011740, 5R01CA133996, HHSN268200782096C; PIs Christopher Amos, Qingyi Wei, Jeffrey E. Lee). For VISP stroke cases of African ancestry, a subset of stroke-free subjects from the Healthy Aging in Neighborhoods of Diversity across the Life Span study (HANDLS) were used as stroke free controls. HANDLS is funded by the National Institute of Aging (1Z01AG000513; PI Michele K. Evans).

Vitamin Intervention for Stroke Prevention (VISP) was funded by the National Institute of Neurological Disorders and Stroke (R01-NS34447). Genome-wide association study data for a subset of VISP participants supported by the National Human Genome Research Institute (U01-HG005160), as part of the Genomics and Randomized Trials Network (PI: Drs Sale and Worrall).

**WHI-OS:** The Women’s Health Initiatives (WHI) program was funded by the National Heart, Lung, and Blood Institute, NIH, US Department of Health and Human Services through contracts N01WH22110, 24152, 32100-2, 32105-6, 32108- 9, 32111-13, 32115, 32118 to 32119, 32122, 42107-26, 42129-32, and 44221. The Hormones and Biomarkers Predicting Stroke (HaBPS) was supported by a grant from the National Institutes of Neurological Disorders and Stroke (R01NS042618).

**WUSTL:** Washington University St. Louis Stroke Study (WUSTL): The collection, extraction of DNA from blood, and storage of specimens were supported by 2 NINDS NIH grants (P50 NS055977 and R01 NS085419. Basic demographic and clinical characterization of stroke phenotype was prospectively collected in the Cognitive Rehabilitation and Recovery Group (CRRG) registry. The Recovery Genomics after Ischemic Stroke (ReGenesIS) study was supported by a grant from the Barnes-Jewish Hospital Foundation.

**Young Lacunar Stroke DNA Resource Plus:** Collection of the UK Young Lacunar Stroke DNA Study (DNA Lacunar) was primarily supported by the Wellcome Trust (WT072952) with additional support from the Stroke Association (TSA 2010/01). Genotyping of the samples, and Dr Traylor, were supported by a Stroke Association Grant (TSA 2013/01).

**Control-only Cohorts:**

**ADHD:** Financial support was received from the “Instituto de Salud Carlos III-FIS", grants PI18/01788, PI19/00721, P19/01224 and PI20/00041, cofinanced by the European Regional Development Fund (ERDF), “Agència de Gestió d’Ajuts Universitaris i de Recerca-AGAUR, Generalitat de Catalunya” (2017SGR1461) and “Departament de Salut”, Government of Catalonia, Spain. Authors wish to thank all participants who kindly participated in this research.

**FINRISK:** Veikko Salomaa was supported by the Finnish Foundation for Cardiovascular Research. Disclosure: Veikko Salomaa has received an honorarium from Sanofi for consulting. He also has ongoing research collaboration with Bayer Ltd. (All unrelated to the present study).

**HCHS/SOL:** The Hispanic Community Health Study/Study of Latinos was carried out as a collaborative study supported by contracts from the National Heart, Lung, and Blood Institute (NHLBI) to the University of North Carolina (N01-HC65233), University of Miami (N01-HC65234), Albert Einstein College of Medicine (N01-HC65235), Northwestern University (N01-HC65236), and San Diego State University (N01- HC65237). The following Institutes/Centers/Offices contribute to the HCHS/SOL through a transfer of funds to the NHLBI: National Center on Minority Health and Health Disparities, the National Institute of Deafness and Other Communications Disorders, the National Institute of Dental and Craniofacial Research, the National Institute of Diabetes and Digestive and Kidney Diseases, the National Institute of Neurological Disorders and Stroke, and the Office of Dietary Supplements

**HRS:** HRS is supported by the National Institute on Aging (NIA U01AG009740). The genotyping was funded as a separate award from the National Institute on Aging (RC2 AG036495). Genotyping was conducted by the NIH Center for Inherited Disease Research (CIDR) at Johns Hopkins University. Genotyping quality control and final preparation of the data were performed by the Genetics Coordinating Center at the University of Washington. HRS genotype data have been deposited in the NIH GWAS repository (dbGaP; accession number: phs000428.v2.p2).

**INMA:** This study was funded by grants from Instituto de Salud Carlos III (CB06/02/0041, G03/176, FIS PI041436, PI081151, PI041705, PI061756, PI091958, and PS09/00432, FIS-FEDER 03/1615, 04/1509, 04/1112, 04/1931 , 05/1079, 05/1052, 06/1213, 07/0314, 09/02647, 11/01007, 11/02591, 11/02038, 13/1944, 13/2032 and CP11/0178), Spanish Ministry of Science and Innovation (SAF2008- 00357), European Commission (ENGAGE project and grant agreement HEALTH-F4- 2007-201413, HEALTH.2010.2.4.5-1, FP7-ENV-2011 cod 282957), Fundació La Marató de TV3, Generalitat de Catalunya-CIRIT 1999SGR 00241 and Conselleria de Sanitat Generalitat Valenciana. Part of the DNA extractions and genotyping was performed at the Spanish National Genotyping Centre (CEGEN-Barcelona). The authors are grateful to Silvia Fochs, Anna Sànchez, Maribel López, Nuria Pey, Muriel Ferrer, Amparo Quiles, Sandra Pérez, Gemma León, Elena Romero, Maria Andreu, Nati Galiana, Maria Dolores Climent, Amparo Cases and Cristina Capo for their 165 assistance in contacting the families and administering the questionnaires. The authors would particularly like to thank all the participants for their generous collaboration. A full roster of the INMA Project Investigators can be found at <https://www.proyectoinma.org/en/inma-project/inma-project-researchers/>

**KORA:** The KORA research platform (KORA, Cooperative Research in the Region of Augsburg) was initiated and financed by the Helmholtz Zentrum München - German Research Center for Environmental Health, which is funded by the German Federal Ministry of Education and Research and by the State of Bavaria. Furthermore, KORA research was supported within the Munich Center of Health Sciences (MC Health), Ludwig-Maximilians-Universität, as part of LMUinnovativ. Funded by the Bavarian State Ministry of Health and Care through the research project DigiMed Bayern

(www.digimed-bayern.de).

**MDC:** The Malmӧ Diet and Cancer Study was supported by the Swedish Research Council (Vetenskapsrådet), Heart and Lung Foundation (Hjärt och Lungfonden), and Swedish Stroke Foundation (Strokeförbundet).

**OAI:** The OAI is a public–private partnership comprised of five contracts (N01-AR-2-2258; N01-AR2-2259; N01-AR-2-2260; N01-AR-2-2261; N01-AR-2-2262) funded by the National Institutes of Health, a branch of the Department of Health and Human Services and conducted by the OAI Study Investigators. Genotyping support was provided by grant RC2-AR-058950 from NIAMS/NIH. Private funding partners include Merck Research Laboratories; Novartis Pharmaceuticals Corporation, GlaxoSmithKline; and Pfizer, Inc. Private sector funding for the OAI is managed by the Foundation for the National Institutes of Health.

**Project MinE/Population based ALS registry, The Netherlands:** The collaboration project is co-funded by the PPP Allowance made available by Health~Holland, Top Sector Life Sciences & Health, to stimulate public-private partnerships. This study was supported by the ALS Foundation Netherlands.
